# Pre-existing cancer cells and induced fibroblasts are key cells for early chemoresistance in ovarian cancer

**DOI:** 10.1101/2024.02.03.24302058

**Authors:** Langyu Gu, Shasha He, Linxiang Wu, Yu Zeng, Yang Zhang, Chenqing Zheng, Chuling Wu, Huishan Xu, Xiaoyan Zhang, Hongwei Shen, Shuzhong Yao, Yufeng Ren, Guofen Yang

## Abstract

Chemoresistance has long been a significant but unresolved issue in the treatment of various cancers, including the most deadly gynecological cancer, the high-grade serous ovary cancer (HGSOC). In this study, single nuclei transcriptome analyses were utilized to identify key cells and core networks for chemoresistance in HGSOC patients with different early responses to platinum-based chemotherapy at the single-cell level. Biomarkers for chemoresistance were also screened using bulk transcriptome data from independent cohorts with larger sample sizes. A total of 62,482 single cells from six samples were analyzed, revealing that chemoresistant cancer cells (Epithelial cells_0) pre-existed within individual patient before treatment. Two network modules formed with hub genes such as hormone-related genes (ESR1 and AR), insulin-related genes (INSR and IGF1R), and CTNNB1, were significantly overexpressed in these cells in the chemoresistant patient. BMP1 and TPM2 could be promise biomarkers in identifying chemoresistant patients before chemotherapy using bulk transcriptome data. Additionally, chemotherapy-induced fibroblasts (Fibroblasts_01_after) emerged as key stromal cells for chemoresistance. One network module containing one subnetwork formed by cholesterol biosynthesis-related genes and one subnetwork formed by cancer-related genes such as STAT3 and MYC, was significantly overexpressed in these cells in the chemoresistant patient. Notably, the NAMPT-INSR was the most prioritized ligand-receptor pair for cells interacting with Fibroblasts_01_after cells and Epithelial cells_0 cells to drive the up-regulation of their core genes, including IL1R1, MYC and INSR itself. Our findings deepen the understandings about mechanisms of early chemoresistance in HGSOC patients.

## Introduction

Chemoresistance presents a formidable obstacle in the effective treatment of various cancers, including the high-grade serous ovarian cancer (HGSOC), which is the most lethal gynecologic malignancy (1). While initial responses to chemotherapy are favorable for most patients (2), approximately one-third of patients responding poorly to chemotherapy, with limited further treatment options available (3). Furthermore, even patients initially responsive well to chemotherapy may eventually develop chemoresistance, contributing to high mortality rates in HGSOC (2). Despite decades of clinical application of chemotherapy, the mechanism behind chemoresistance remains largely unknown. Although individual chemoresistant targets have been identified using classical biological experimental methods, their efficacy is usually limited due to the inherent complexity of biological organisms. We thus need to analyze the mechanism of chemoresistance at a higher resolution and at the systemic network level.

Cell populations are fundamental to the execution of biological functions in humans, and chemoresistance represents a selection process favoring cell populations with adaptive fitness phenotypes. A critical question arises: Do different responses to chemotherapy pre-exist in patients before treatment, or are they acquired during treatment? Answering these questions necessitates a single-cell level analysis, including the differentiation of cancer cells with varying chemotherapy responses and the interactions among cells in the tumor microenvironments of patients with different responses. Previous studies in different cancer types, such as breast cancer, rectal cancer, and oral squamous cell carcinomas, have revealed diverse mechanisms of chemoresistance, with either showing pre-existing genetic mutations for chemoresistance (4), or demonstrating acquired transcriptional profile reprogramming during treatment (5), or both (6). These findings underscore the need for specific analyses tailored to each cancer type. One recent study has provided valuable insights into the mechanisms of chemoresistance in HGSOC, but it focused on metastatic sites (7), leaving the mechanisms at the primary sites of HGSOC relatively unclear. Furthermore, the use of platinum-free intervals to characterize chemoresistance remains controversial and may not directly reflect chemoresistance (8).

Therefore, in this study, we sought to investigate and compare cellular composition and corresponding transcriptional profiles at the primary sites before and after platinum-based chemotherapy in patients with varying responses at the single-cell level. Our objectives were to address the following questions: Which cell types are key cells for early chemoresistance? Are the corresponding gene co-expression networks pre-existing or acquired? What are specific cellular interactions in the microenvironments in chemoresistant patients? Our findings will deepen our understandings about mechanisms of chemoresistance, and can be helpful for finding potential biomarkers and therapeurtic targets for early chemoresistant HGSOC patients.

## Materials and Methods

### Sampling and Library Construction

This study was approved by the Ethics Committee for Clinical Research and Animal Trials of the First Affiliated Hospital of Sun Yat-sen University (ethics approval No. 2021726). Six fresh paired primary HGSOC samples before and after chemotherapy were collected from three patients who received neoadjuvant chemotherapy (NACT) between 2021 and 2022 at the First Affiliated Hospital, Sun Yat-sen University in Guangzhou, China. Clinical information can be found in the Supplementary File 1. The chemotherapy regimen consisted of three cycles of paclitaxel plus carboplatin. Paired samples were obtained during laparoscopy and interval debulking surgery before and after chemotherapy. Fresh tissues were immediately frozen in liquid nitrogen and transported by dry ice. Single nuclei transcriptome libraries construction and sequencing were conducted by Novogene (China) following 10x Genomics instructions (https://support.10xgenomics.com). Briefly, libraries were constructed using the Chromium Next GEM Chip G Single Cell Kit, 48 rxns (PN-100020), and Chromium Next GEM Single Cell 3’ GEM, Library & Gel Bead Kit v3.1, 16 rxns PN-1000121. Libraries with PE150 (paired-end reads, 150bp) were sequenced on an Illumina NovaSeq 6000.

### Chemotherapy Response Evaluation

All patients underwent baseline 18F-FDG PET/CT before and after chemotherapy (Figure 1A). 18F-FDG PET/CT was performed after at least 6 hours of fasting and with a glucose level lower than 10 mmol/L. The PET/CT scan coverage ranged from the top of the head to the mid-thigh. The ordered-subset expectation maximization iterative reconstruction method was used to reconstruct the data. The total metabolic tumor volume (tMTV) and total lesion glycolysis (TLG) were measured and calculated. Based on PERCIST 1.0 (9), we differentiated patients as partial responses to chemotherapy (PR1 and PR2) and progress disease (PD) (we named these IDs for this study and they were not used to identify patients outside the research group) (Figure 1B).

**Figure 1.**
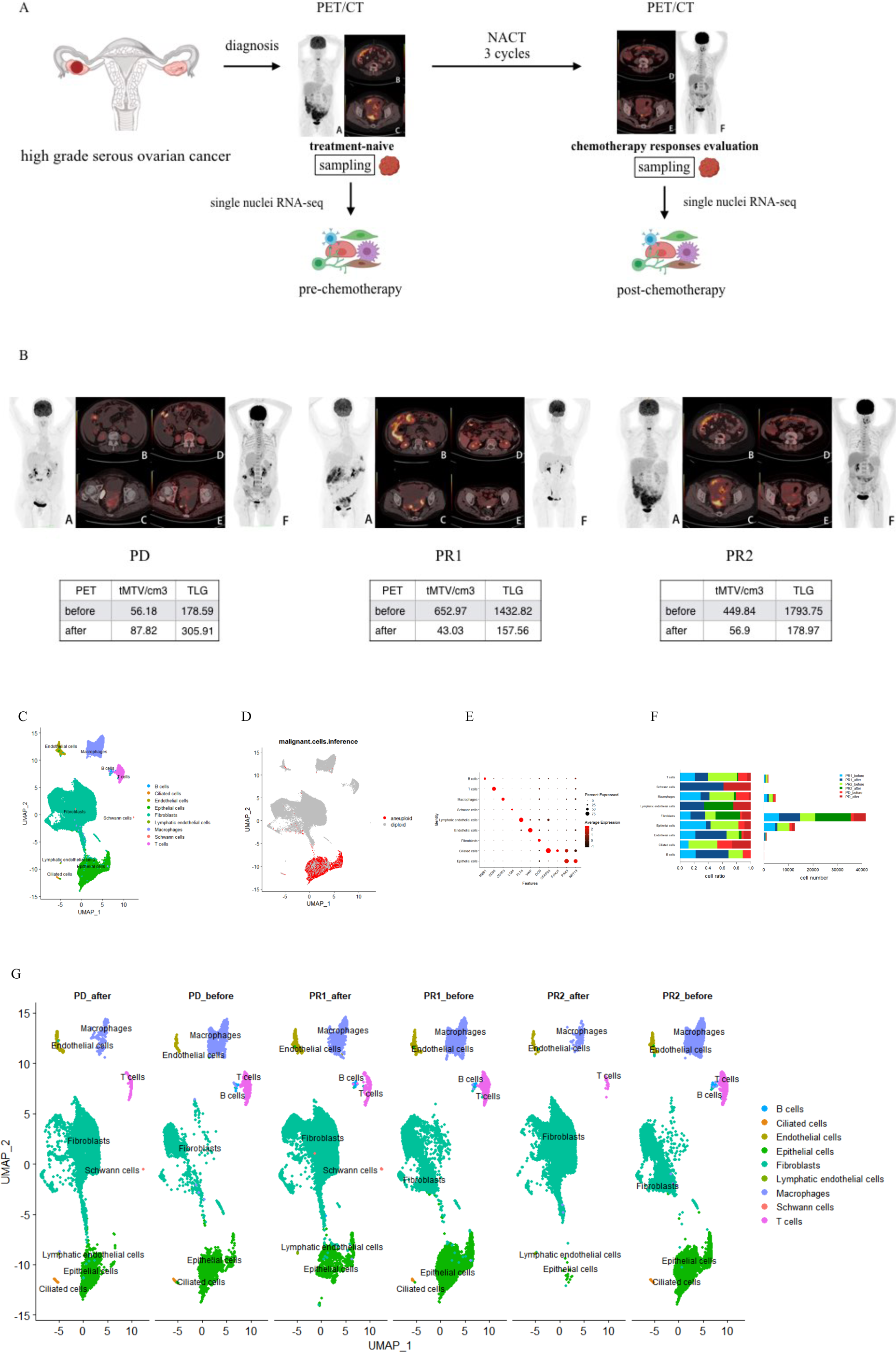
Schematic of sampling and analysis results. (A) Schematic of sampling. Fresh paired primary samples before and after chemotherapy were collected from three patients who received neoadjuvant chemotherapy (NACT). The chemotherapy regimen consisted of three cycles of paclitaxel plus carboplatin. PET/CT was used to evaluate chemotherapy responses before and after chemotherapy for each patient. Paired samples were collected during laparoscopy and interval debulking surgery before and after chemotherapy. Single nuclei transcriptome libraries constructions were conducted following 10 x Genomics instructions. (B) Chemotherapy responses evaluation. tMTV (total metabolic tumor volume) and TLG (total lesion glycolysis) were used to evaluate chemotherapy responses. PR1 represents patient 1 with a partial response to chemotherapy, PR2 represents patient 2 with a partial response to chemotherapy, and PD represents progressive disease (chemoresistant patient). (C) The UMAPT plot of cell atlas of integrated samples showing major cell clusters. (D) Malignant cells inference. Aneuploid cells (malignant cells) and diploid cells (non-malignant cells) were identified. (E) The dot plot of marker genes expression for each major cell cluster. (F) Cell numbers and cell ratios of each major cell cluster in each individual sample. (G) The UMAP plot of cell atlas showing major cell clusters in each individual sample among patients before and after chemotherapy. PD_after represents the post-chemotherapy sample of the chemoresistant patient, PD_before represents the pre-chemotherapy sample of the chemoresistant patient, PR1_after represents the post-chemotherapy sample of patient 1 with partial response to chemotherapy, PR1_before represents the pre-chemotherapy sample of patient 1 with partial response to chemotherapy, PR2_after represents the post-chemotherapy sample of patient 2 with partial response to chemotherapy, and PR2_before represents the pre-chemotherapy sample of patient 2 with partial response to chemotherapy.

### Data Pre-processing

Raw data were analyzed with 10x Genomics Cell Ranger 6.1.1. The GRCh38 Ensembl build genome (refdata-gex-GRCh38-2020-A) was used as the reference. The filtered feature barcode matrix was used for further data analysis. The Seurat v4 R package was used for data pre-processing (10). Only genes expressed in at least three cells, and cells with at least 200 unique Molecular Identifiers (UMI) were retained. Quality control metrics were plotted for individual samples and used for filtering cells with aberrantly high UMI, which could be originated from cell doublets or multiplets. The PercentageFeatureSet() function was used to calculate the percentage of counts originating from mitochondrial DNA (mtDNA). Low-quality or dead cells with abnormal mitochondrial contamination were removed. Cell clusters with fewer than 10 cells were also removed. Cell cycle effects were evaluated with CellCycleScoring() function. Doublets were assessed using the DoubletFinder v2.0 R package (11).

Based on the plotting results (Supplementary File 2), quality control was performed for each sample. For PR1_before (pre-chemotherapy sample of patient 1 with partial response to chemotherapy), cells with 200 < UMI < 7500, mtDNA percentage < 8, ribosome percentage < 4 were retained, and 984 doublets were removed. Similarly, for PR1_after (post-chemotherapy sample of patient 1 with partial response to chemotherapy), cells with 200 < UMI < 5000, mtDNA percentage < 2, ribosome percentage < 1 were retained, and 831 doublets were removed. For PR2_before (pre-chemotherapy sample of patient 2 with partial response to chemotherapy), cells with 200 < UMI < 7500, mtDNA counts < 6%, ribosome percentage < 2 were retained, and 1057 doublets were removed. For PR2_after (post-chemotherapy sample of patient 2 with partial response to chemotherapy), cells with 200 < UMI < 4300, mtDNA percentage < 2.5%, ribosome percentage < 2 were retained, and 1119 doublets were removed. For PD_before (pre-chemotherapy sample of the chemoresistant patient), cells with 200 < UMI < 6000, mtDNA percentage < 10, ribosome percentage < 2 were retained, and 144 doublets were removed. For PD_after (post-chemotherapy sample of the chemoresistant patient), cells with 200 < UMI < 6000, mtDNA percentage < 1, ribosome percentage < 2 were retained, and 367 doublets were removed.

### Data Integration, Clustering, and Differential Gene Expression Analysis

The preprocessed data were used for data integration analysis using Seurat to correct batch effects among samples (10). 2000 highly variable expressed genes detected for each sample were used to find anchors for data integration. Subsequently, the integrated data was scaled, and the principal components were computed. The top 30 principal components were identified using an elbow plot. Clustering identification was performed at a proper resolution that can differentiate major cell types. The UMAP dimensionality reduction was used for visualization. Diffential expressed (DE) genes were identified with a significance threshold of adjusted *p* value < 0.05 and log2FoldChange > 0.5. KEGG pathways enrichment analyses were performed using the STRING database (https://string-db.org/) (12).

### Malignant Cells Inference

Malignant cells were inferred based on genomic copy number variation (CNV) using CopyKAT v1.1.0 (13). Cells with extensive genome-wide copy number aberrations (aneuploidy) were considered as malignant tumor cells. Parameters were set as ngene.chr=10, win.size=25, KS.cut=0.1, distance=“euclidean”. Only cells that met the criteria were presented. For the PR2_after which contained only a small number of epithelial cells (15 cells), we assumed that all these epithelial cells were malignant cells, since they all overexpressed malignance cell markers.

### Gene Co-expression Network Construction

hdWGCNA (14) was used to construct high dimensional gene co-expression network modules for target cell clusters. Genes expressed in at least 5% of cells were used for analyses. Batch effects were corrected during analyses by setting group.by and group.by.vars parameters. The number of cells to be aggregated for metacells construction was 25, and the max_shared was set as 10 as default. The TestSoftPowers was used to determine a proper value of the soft power threshold. The implemented AddModuleScore function was applied to compute hub genes signature scores for each module. Differential module eigengene analysis was performed between target cell groups. Hub genes overexpressed in the target cells in the network modules were further used to construct protein-protein network using the STRING database (12). The thickness of network edges represents the strength of data support. The minimum required interaction score was set as 0.7 (high confidence). Only genes with interaction degrees >1 were presented.

### Survival Analyses

Survival analyses were conducted using three publicly available bulk transcriptome datasets with clinical information, including GSE102073 (n=70), PRJNA866991 (n=41), and GSE32062 (n=260). Kaplan-Meier (K-M) survival curves were constructed using the survival 3.5 R package (15).

### Bulk Transcriptome Analyses

For the GSE102073 cohort, the normalized gene expression matrix was obtained directly from the GEO database. For the PRJNA866991 cohort, raw reads were retrieved from the NCBI database and aligned to the GRCh38 Ensembl build human genome using HISAT2 v2.1.0 (16). SAMTOOLs v1.7 (17,18) was utilized to convert sam files to bam files, sort and index bam files, and obtain the final counts. Featurecounts v2.0.4 (19) was used to obtain the counts at the gene level. The DESeq2 v1.38.3 (20) was employed to normalize the count files and generate the final expression matrix. For the GSE32062 cohort, CEL files were first downloaded from the GEO database. The affy v1.74 R package (21) was used to process the CEL files, and rma was applied for normalization. The hgu133plus.db was used to extract the gene expression matrix at the gene level.

### Cell-Cell Interaction Analyses

Nichenetr (22) was used to identify prioritized ligand-receptor-target genes involved in the interactions from senders to receivers under different conditions. All genes expressed in at least 10% of receiving cells were used as the background geneset.

### RNA Velocity Analyses

RNA velocity (the change in mRNA abundance calculated by relating the abundance of unspliced and spliced mRNA) was estimated using Scvelo (23) following the instructions (https://scvelo.readthedocs.io/en/stable/). The stochastic model was chosen to account for stochasticity in gene expression, and the combination of velocities across genes was used to estimate the future state of individual cells.

### Data Availability

All data produced in the present study are available upon reasonable request to the authors.

## Results

### Cell atlas before and after chemotherapy in patients with different responses

We collected six samples at primary ovary sites from three HGSOC patients paired before and after chemotherapy. The responses to chemotherapy were evaluated using PET/CT (Figure 1A and Figure 1B). The results of tMTV and TLG of each patient before and after chemotherapy were given in Figure 1B. Based on PERCIST 1.0 (9), we differentiated patients as partial responses to chemotherapy (PR1 and PR2) and progress disease (PD). After quality control, a total of 62482 single cells were grouped into nine major clusters, including 12668 epithelial cells (KRT19^+^), 41522 fibroblasts (DCN^+^), 1825 T cells (CD96^+^), 4850 Macrophages (CD163^+^), 1120 Endothelial cells (VWF^+^), 75 Lymphatic endothelial cells (FLT4^+^), 213 B cells (MZB1^+^), 43 Schwann cells (LGI4^+^), and 166 Ciliated-secretory intermediate cells (CFAP54^+^FOXJ1^+^PAX8^+^) (24,25) (Figure 1C). Malignant cells inference revealed that malignant cells were mainly epithelial cells (Figure 1D), which is consistent with the pathological characteristics of HGSOC as an epithelial cancer. A small portion of ciliated-secretory intermediate cells were also identified as malignant cells, which can be explained since they were assumed to be the main origin of HGSOC (24,25). Expression profiles of corresponding markers in each cell type were provided in Figure 1E. The cell numbers and ratios in each major cell cluster from individual patients were also presented in Figure 1F. The distribution of major cell clusters in each sample among patients before and after chemotherapy were illustrated in Figure 1G.

### Chemoresistant cancer cells were pre-existing in the PD patient

Five subclusters of epithelial cells were identified in total (Figure 2A and 2B). No genes were specifically overexpressed in Epithelial cells_0, indicating that this subcluster represented a common cancer cell type. Epithelial cells_1 specifically overexpressed MKI67 and enriched cell proliferation-related pathways, such as homologous recombination, DNA replication, and the cell cycle. Epithelial cells_2 specifically overexpressed the hypoxia-related gene PFKFB. Epithelial cells_3 specifically overexpressed DOCK8 and PTPRC, and enriched immune response-related pathways, such as the B cell receptor and T cell receptor pathways. Epithelial cells_4, which only appeared in the partial response after chemotherapy, specifically overexpressed DACH2 and OVGP1, and enriched cancer-related pathways (Figure 2C, 2D).

**Figure 2.**
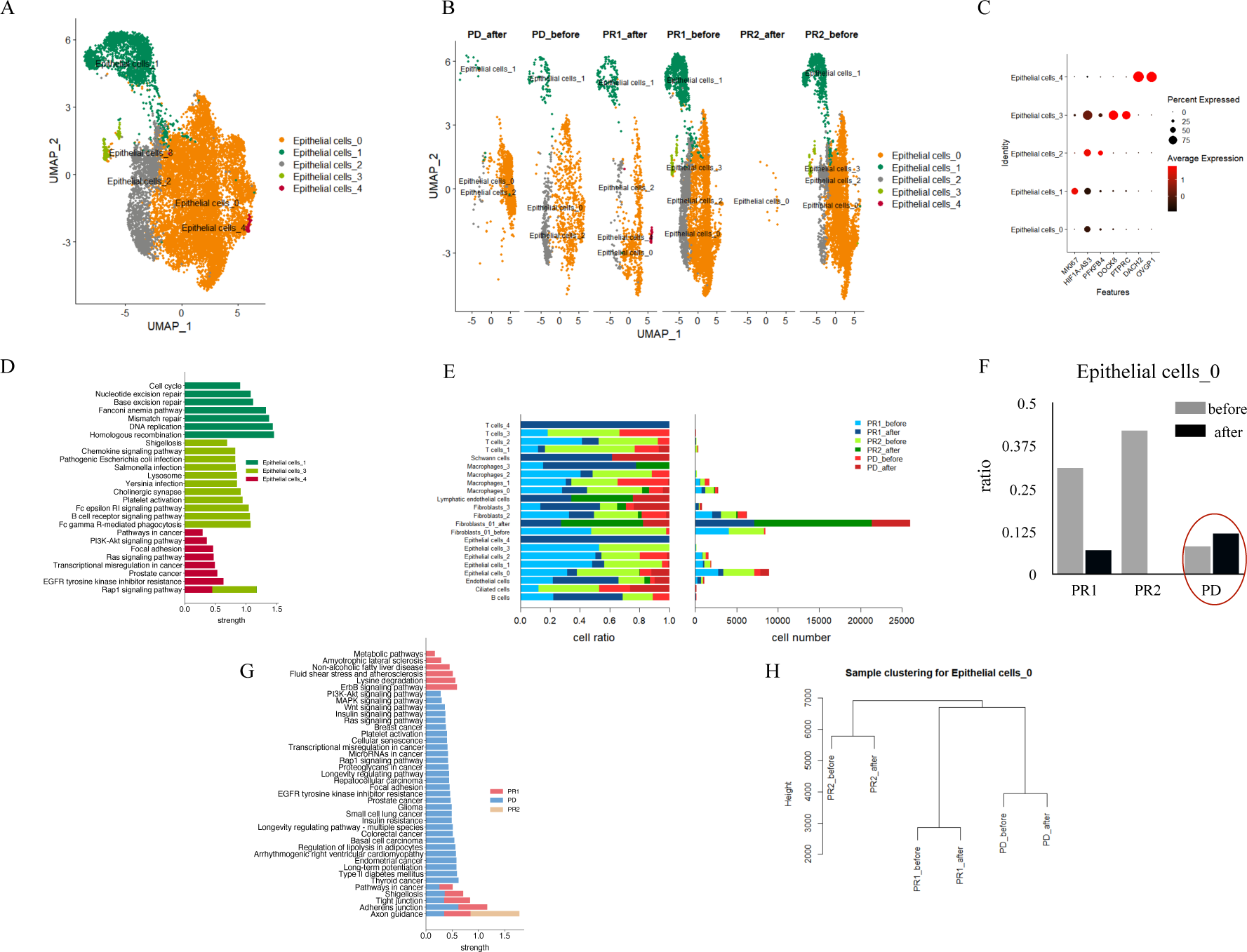
(A) The UMAP plot of cell atlas for five epithelial cells subclusters. (B) The UMAP plot of cell atlas showing five epithelial cells subclusters in each individual sample among patients before and after chemotherapy. PD_after represents the post-chemotherapy sample of the chemoresistant patient, PD_before represents the pre-chemotherapy sample of the chemoresistant patient, PR1_after represents the post-chemotherapy sample of patient 1 with a partial response to chemotherapy, PR1_before represents the pre-chemotherapy sample of patient 1 with a partial response to chemotherapy, PR2_after represents the post-chemotherapy sample of patient 2 with a partial response to chemotherapy, PR2_before represents the pre-chemotherapy sample of patient 2 with a partial response to chemotherapy. (C) The dot plot of marker genes expression for epithelial cells subclusters. (D) KEGG pathway enrichment for epithelial cells subclusters. (E) Cell numbers and cell ratios of cell subclusters in each individual sample. (F) Cell ratio comparisons of the Epithelial cells_0 subcluster between samples before and after chemotherapy in each patient. (G) KEGG pathway enrichment for the Epithelial cells_0 subcluster among samples after chemotherapy. (H) Sample clustering using cell average transcriptional profiles clustered Epithelial cells_0 cells based on individual patient ID rather than treatment condition (before or after chemotherapy).

Cell proportion analysis showed that Epithelial cells_0 cells were highly enriched in the PD_after, indicating its chemoresistant character (Figure 2E and 2F). DE genes retrieved from comparisons among samples after chemotherapy for each subcluster also revealed that Epithelial cells_0 cells enriched many cancer-related pathways in the PD_after, such as the Wnt signaling pathway, the MAPK signaling pathway, the insulin signaling pathway, the cellular senescence pathway, and the longevity regulating pathway (Figure 2G). In contrast, although other subclusters also remained after chemotherapy in the PD_after, their proportions were largely reduced, and no cancer-related pathways were enriched, indicating their fragility to chemotherapy. Thus, we proposed that Epithelial cells_0 cells were the key cancer cells for early chemotherapy responses. It is worth noting that clustering analysis using cell average transcriptional profiles clustered Epithelial cells_0 cells based on individual patient ID rather than treatment condition (before or after chemotherapy) (Figure 2H), indicating that transcriptional profiles for chemotherapy responses were largely pre-existing in Epithelial cells_0 cells in individual patients before chemotherapy.

Gene co-expression networks construction further supported the above results. We identified six co-expression modules for Epithelial cells_0 cells in total. Module eigengene (ME) represents the first principal component of the gene expression matrix comprising each module, and was thus used to summarize the gene expression profile of the entire module. Figure 3A presented the summarization of the expression of each module obtained by calculating gene signature scores from MEs for the top 25 hub genes for each module. To correct batch effects, harmony batch correction was applied to produce harmonized MEs (hMEs) (Figure 3B). We can see that, again, pre- and post-chemotherapy samples from the same patient expressed similar modules (Figure 3B). For example, the PD patient overexpressed M3 and M4 modules both before and after chemotherapy. Differential module eigengene analysis revealed that no module was significantlly overexpressed in the post-chemotherapy sample when compared to the pre-chemotherapy sample from the same patient. For example, although the adjust *p* value of M3 in the PD_after was significant when compared to PD_before, its foldchange was small (Figure 3C left). Although the adjust *p* values of M3 and M4 modules were significant in the PR1_after when compared to the ones in the PR1_before, they actually expressed little in theses samples (Figure 3C middle). This can be also applied to M4 and M6 modules in the PR2 patient (Figure 3C right). These results confirmed again that the transcriptional profiles for different chemotherapy responses were pre-existing in cancer cells before chemotherapy.

**Figure 3.**
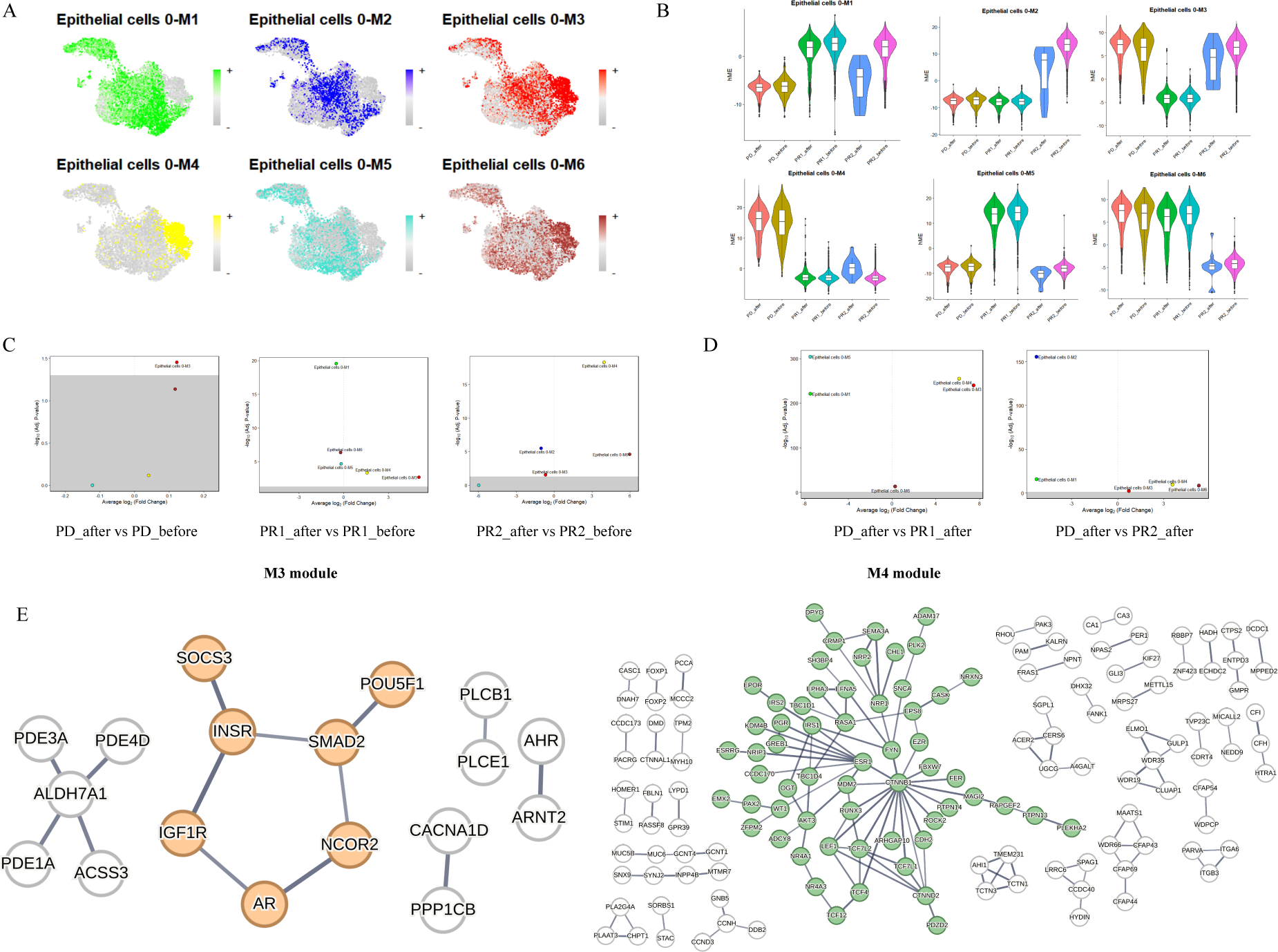
Gene co-expression networks construction and comparisons for Epithelial cells_0. (A) The UMAP plots of the hub gene signature score for each module constructed for Epithelial cells_0. (B) Violin plots of the harmonized module eigengenes (hMEs) in each sample for each module. (C) Differential module eigengene comparisons between post- and pre-samples within each individual patient. (D) Differential module eigengene comparisons between the PD_after and the PR1_after or the PR2_after, respectively. (E) Co-expression networks visualization of Module 3 and Module 4. PD_after: post-chemotherapy sample of the chemoresistant patient. PD_before: pre-chemotherapy sample of the chemoresistant patient. PR1_after: post-chemotherapy sample of the patient 1 with partial response to chemotherapy. PR1_before: pre-chemotherapy of the patient 1 with partial response to chemotherapy. PR2_after: post-chemotherapy sample of the patient 2 with partial response to chemotherapy. PR2_before: pre-chemotherapy of the patient 2 with partial response to chemotherapy.

Notably, when comparing Epithelial cells_0 cells among samples after chemotherapy, M3 and M4 modules were significantly overexpressed in Epithelial cells_0 cells in the PD_after when compared to the ones in either PR1_after or PR2_after (Figure 3D), indicating the chemoresistant characters of these two modules. Further protein-protein co-expression networks using these module genes overexpressed in the PD_after showed that insulin-related receptors (INSR and IGF1R), hormone receptor (AR), NCOR2, and SMAD2 formed a tight connective network in module M3, and a tight network with CTNNB1and ESR1 as the core hubs in module M4 (Figure 3E).

The presence of pre-existing chemoresistance transcriptional profiles in cancer cells before chemotherapy lays the foundation for the search for relevant pre-treatment biomarkers. Survival analyses were thus conducted using publicly available bulk transcriptome data with progression-free survival (PFS) information, representing the time to first recurrence after chemotherapy. These analyses were performed using datasets from three independent cohorts: PRJNA866991 (n=41), GSE32062 (n=260), and GSE102073 (n=70). It is important to note that these cohorts utilized bulk transcriptome data, where gene expression represents the expression level of the entire sample rather than a specific cell group. Additionally, these data were only retrieved from pre-chemotherapy conditions. We thus first identified DE genes that were significantly overexpressed in the whole sample of PD_before when compared to PR1_before and PR2_before. Subsequent survival analyses of these genes revealed that only two genes, BMP1 and TPM2, were significantly associated with low PFS across all three cohorts (Figure 4A). Both genes were also significantly overexpressed in Epithelial cells_0 cells in the PD patient when compared to samples of PR patients (Figure 4B).

**Figure 4.**
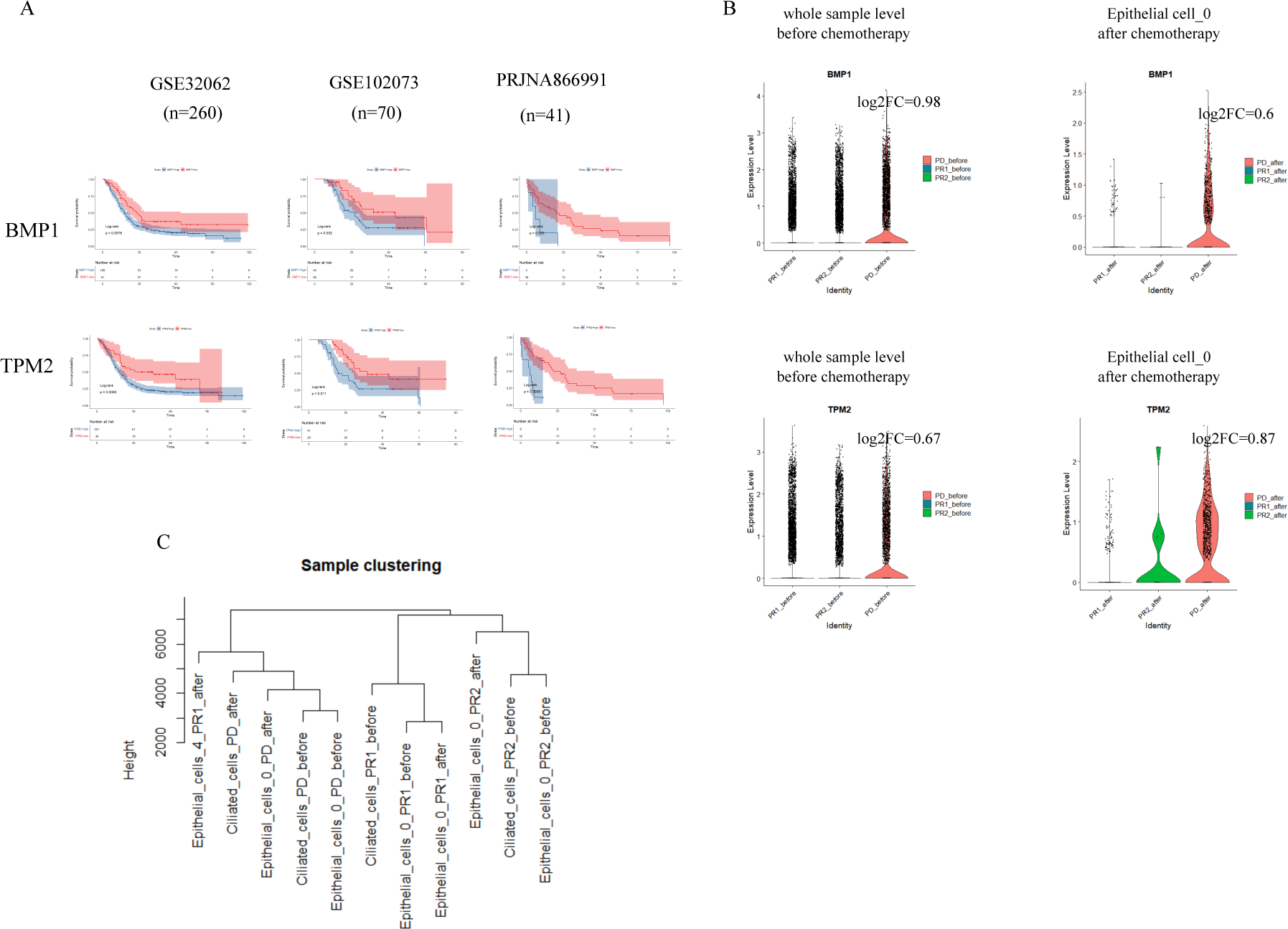
(A) Kaplan-Meier survival curves on progression-free survival (PFS) for genes BMP1 and TPM2 from three independent cohorts. (B) Violin plots showing gene expression patterns among samples at the sample level before chemotherapy and at the Epithelial cells_0 level after chemotherapy. (C) Sample clustering revealing that the expression profiles of Ciliated-secretory intermediate cells were clustered based on individual patients rather than treatment condition.

We also investigated chemoresistant genes in ciliated-secretory intermediate cells since they were also remained in the PD_after, and are assumed to be the main origin of the high-grade serous ovarian carcinoma (HGSOC) (24,25). Similar to Epithelial cells_0 cells, sample clustering also revealed that the expression profiles of ciliated-secretory intermediate cells were clustered based on individual patients rather than treatment condition (Figure 4C). There were only 17 genes overexpressed in Epithelial cells_0 cells when compared to ciliated-secretory intermediate cells in the PD_after with no pathways enriched, indicating their similar transcriptional profiles. This indicated that ciliated-secretory intermediate cells in the PD_after could also be chemoresistant, and the corresponding transcriptional profiles were also pre-existing in individual patients before treatment.

### The induced fibroblasts after chemothrerapy are key cells for chemoresistance

We initially identified four fibroblasts subclusters (Figure 5A). Fibroblasts_0 occupied a large proportion in the tumor microenvironment after chemotherapy, indicating its association with chemotherapy. Notably, Fibroblasts_0 was difficult to be clearly separated from Fibroblasts_1 in the UMAP plot with different resolutions, indicating that Fibroblasts_0 cells were induced from Fibroblasts_1 during the treatment. Transcriptional dynamics analysis also revealed that the transcriptional dynamics direction of Fibroblasts_1 before chemotherapy changed to the same direction of Fibroblasts_0 after chemotherapy, except for a small portion of cells whose direction was more inclined towards the pre-treatment direction. However, no DE gene was specifically found to be overexpressed in this part. Additionally, when we conducted sampling clustering using cellular average transcriptional profiles, we found that unlike other subclusters, Fibroblasts_0 was always clustered together with Fibroblasts_1 in each sample (Figure 5B and 5C). We thus treated Fibroblasts_0 and Fibroblasts_1 as one cluster named Fibroblasts_01, and differentiated pre-chemotherapy and post-chemotherapy clusters into Fibroblasts_01_before and Fibroblasts_01_after (Figure 5D, Figure 5E and 5F).

**Figure 5.**
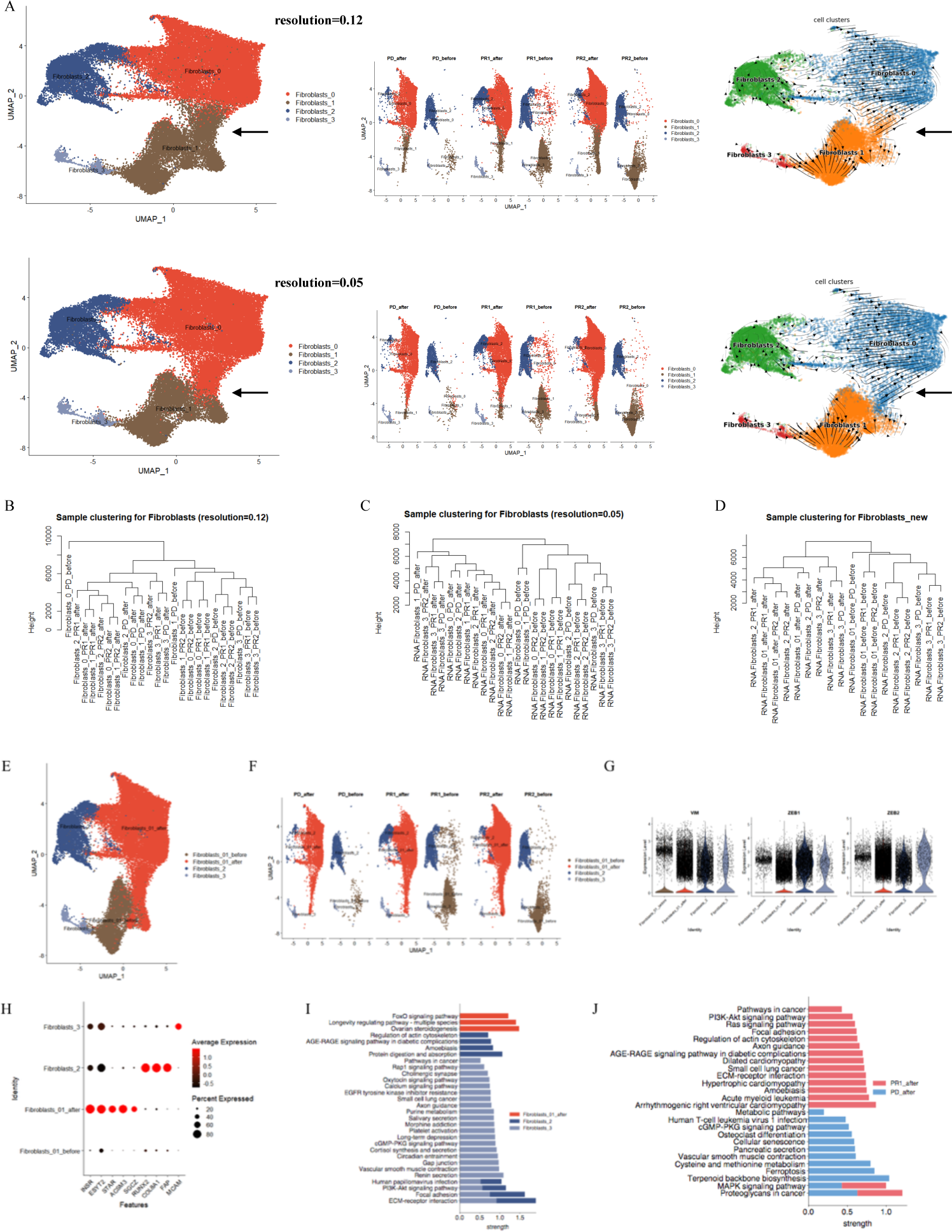
(A) The UMAP plot of cell atlas for fibroblasts subclusters with different resolutions. Left: the UMAP plot of cell atlas for fibroblasts subclusters in integrated samples. Middle: the UMAP plot of cell atlas for fibroblasts subclusters in each sample among patients before and after chemotherapy. Right: transcriptional dynamic depicted by RNA velocity for each subcluster. (B) Sampling clustering using cellular average transcriptional profiles for fibroblasts subclusters under resolution 0.12. (C) Sampling clustering using cellular average transcriptional profiles for fibroblasts subclusters under resolution 0.05. (D) Sampling clustering using cellular average transcriptional profiles for fibroblasts subclusters after treating Fibroblasts_0 and Fibroblasts_1 as one subcluster, and differentiated pre-chemotherapy and post-chemotherapy clusters into Fibroblasts_01_before and Fibroblasts_01_after. (E) UMAP plot of fibroblast subclusters in integrated samples. (F) UMAP plot of fibroblast subclusters in each sample from patients before and after chemotherapy. (G) Violin plots depicting the expression of epithelial-mesenchymal transition (EMT) markers in each subcluster. (H) Dot plot showing marker gene expression in fibroblast subclusters. (I) KEGG pathway enrichment analysis for fibroblast subclusters. (J) KEGG pathway enrichment for overexpressed genes in the PD_after for each subcluster. PD_after: Post-chemotherapy sample of the chemoresistant patient. PD_before: Pre-chemotherapy sample of the chemoresistant patient. PR1_after: Post-chemotherapy sample of patient 1 with partial response to chemotherapy. PR1_before: Pre-chemotherapy sample of patient 1 with partial response to chemotherapy. PR2_after: Post-chemotherapy sample of patient 2 with partial response to chemotherapy. PR2_before: Pre-chemotherapy sample of patient 2 with partial response to chemotherapy.

All identified fibroblasts were epithelial-mesenchymal transition (EMT) fibroblasts, as evidenced by their high expression of corresponding markers VIM, ZEB1, and ZEB2 (Figure 5G). Although no genes were specifically overexpressed in Fibroblasts_01_before, several genes were specifically overexpressed in Fibroblasts_01_after after chemotherapy. The top five overexpressed DE genes for Fibroblasts_01_after in the PD_after were SGCZ, ACSM3, STAR, ESYT2, and INSR. Three pathways were enriched for this subcluster, including ovarian steroidogenesis, longevity regulating pathway-multiple species, and FoxO signaling pathway. The corresponding genes were IGF1R, INSR, STAR, FOXO1, and SGK1. Additionally, Fibroblasts_2 (FAP^+^ COL8A1^+^ RUNX2^+^) specifically overexpressed FAP, COL8A1, and RUNX2, and enrichment in pathways related to ovarian steroidogenesis, regulation of actin cytoskeleton, AGE-RAGE signaling pathway in diabetic complications, and amoebiasis. Fibroblasts_3 (MCAM^+^) specifically overexpressed MCAM and enriched pathways such as pathways in cancer, cortisol synthesis and secretion, vascular smooth muscle contraction, and cGMP-PKG signaling pathway (Figure 5H and 5I).

The sample clustering tree using average transcriptional profiles showed that all subclusters were clustered by treatment condition rather than patient IDs (Figure 5D), signifying that treatment condition played a critical role in fibroblasts among patients. When examining the enrichment of overexpressed genes in the PD_after compared to PR1_after and PR2_after for each subcluster, only overexpressed genes in the Fibroblasts_01_after enriched many cancer-related signaling pathways, indicating that Fibroblasts_01_after in the PD_after exhibited a high degree of malignancy (Figure 5J). Combined with the abundance after treatment for Fibroblasts_01_after (Figure 5A), we thus proposed that Fibroblasts_01_after cells were the key cells for chemotherapy responses.

Gene co-expression networks construction revealed five network modules for Fibroblasts_01_after cells in total (Figure 6A and 6B). The M2 module was significantly overexpressed in the PD_after when compared to either PR1_after or PR2_after, indicating its chemoresistance characters (Figure 6C). Protein-protein network using hub genes of this module that overexpressed in the PD_after revealed two subnetworks. One was formed by many cholesterol biosynthesis-related genes, such as HMGCR, SREBF2, MSMO1, MVD, ACAT2 and INSIG1. The other subnetwork was formed by well-known cancer promoting genes such as MYC and STAT3 (Figure 6D), and enriched in many cancer-related pathways such as HIF-1 signaling pathway, insulin resistance, JAK-STAT signalling pathway, and type II diabetes mellitus.

**Figure 6.**
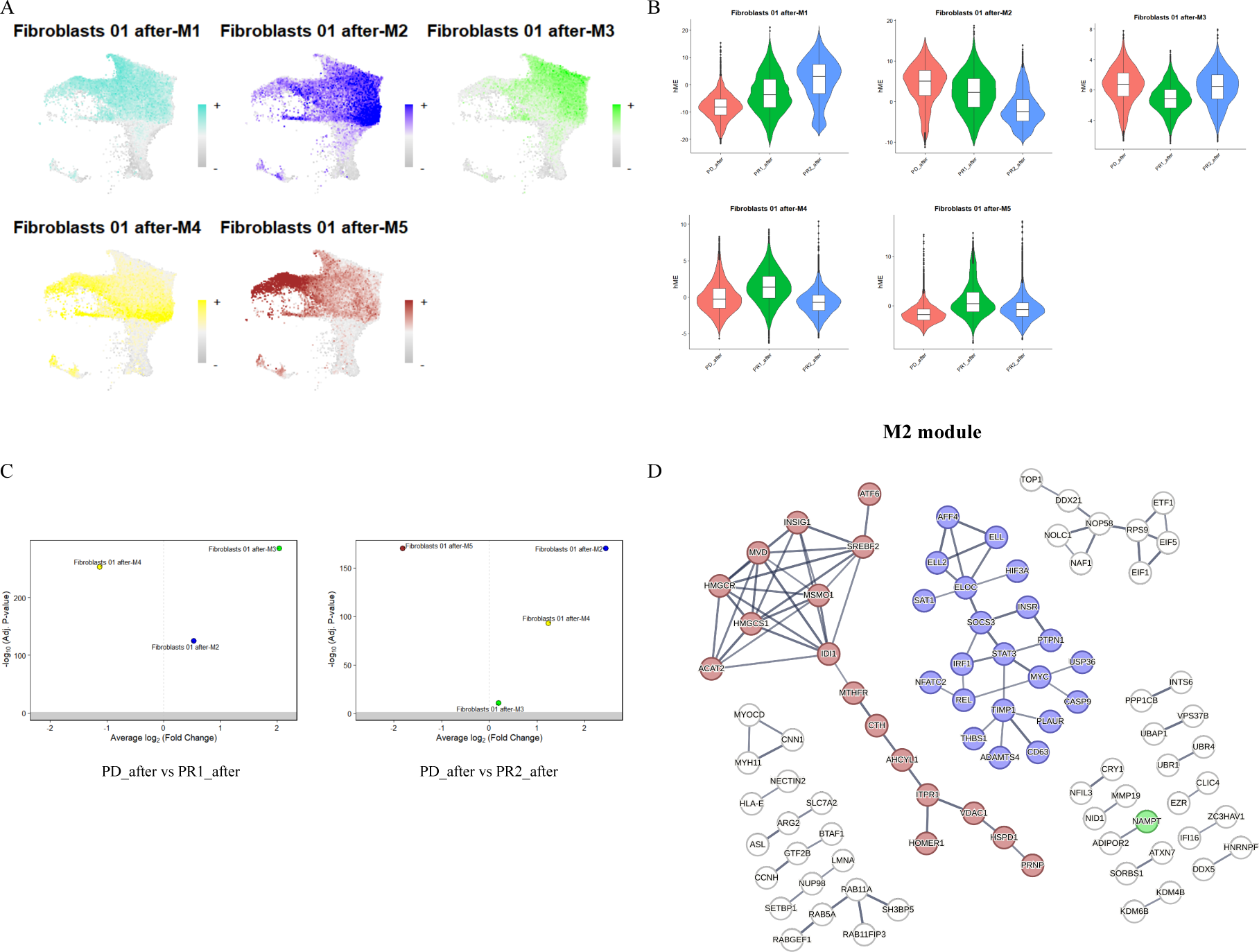
Gene co-expression networks construction and comparisons for Fibroblasts_01_after. (A) The UMAP plots of the hub gene signature score of five modules constructed for Epithelial cells_0. (B) Violin plots of the harmonized module eigengenes (hMEs) in each post-treatment sample for each module. (D) Differential module eigengene comparisons between PD_after and PR1_after or PR2_after, respectively. (E) Co-expression network visualization using hub genes of Module 2. PD_after: post-chemotherapy sample of the chemoresistant patient. PR1_after: post-chemotherapy sample of the patient 1 with partial response to chemotherapy. PR2_after: post-chemotherapy sample of the patient 2 with partial response to chemotherapy.

### Other stromal cells and immune cells in the microenvironment were generally tumour-promoting among patients after chemotherapy

We identified four macrophage subclusters (Figure 7), including CD163^+^ M2 macrophages (Macrophages_0), hypoxic macrophages (Macrophages_1) which overexpressed hypoxia-related genes (HK2^+^) (26), RRM2^+^ M2 macrophages (Macrophages_2), and fibrosis-like macrophages (Macrophages_3) that overexpressed fibroblast-related genes (PDGFRA^+^ and CALD1^+^), consistent with previous reports of macrophages being capable of transdifferentiating into myofibroblasts (27). Following chemotherapy, a large proportion of Macrophages_0 cells and a few Macrophages_1 cells remained in the PD_after. Only 31 genes were found to be overexpressed in the PD_after compared to PR1_after and PR2_after for Macrophages_0, and no genes were overexpressed in the PD_after for Macrophages_1.

**Figure 7.**
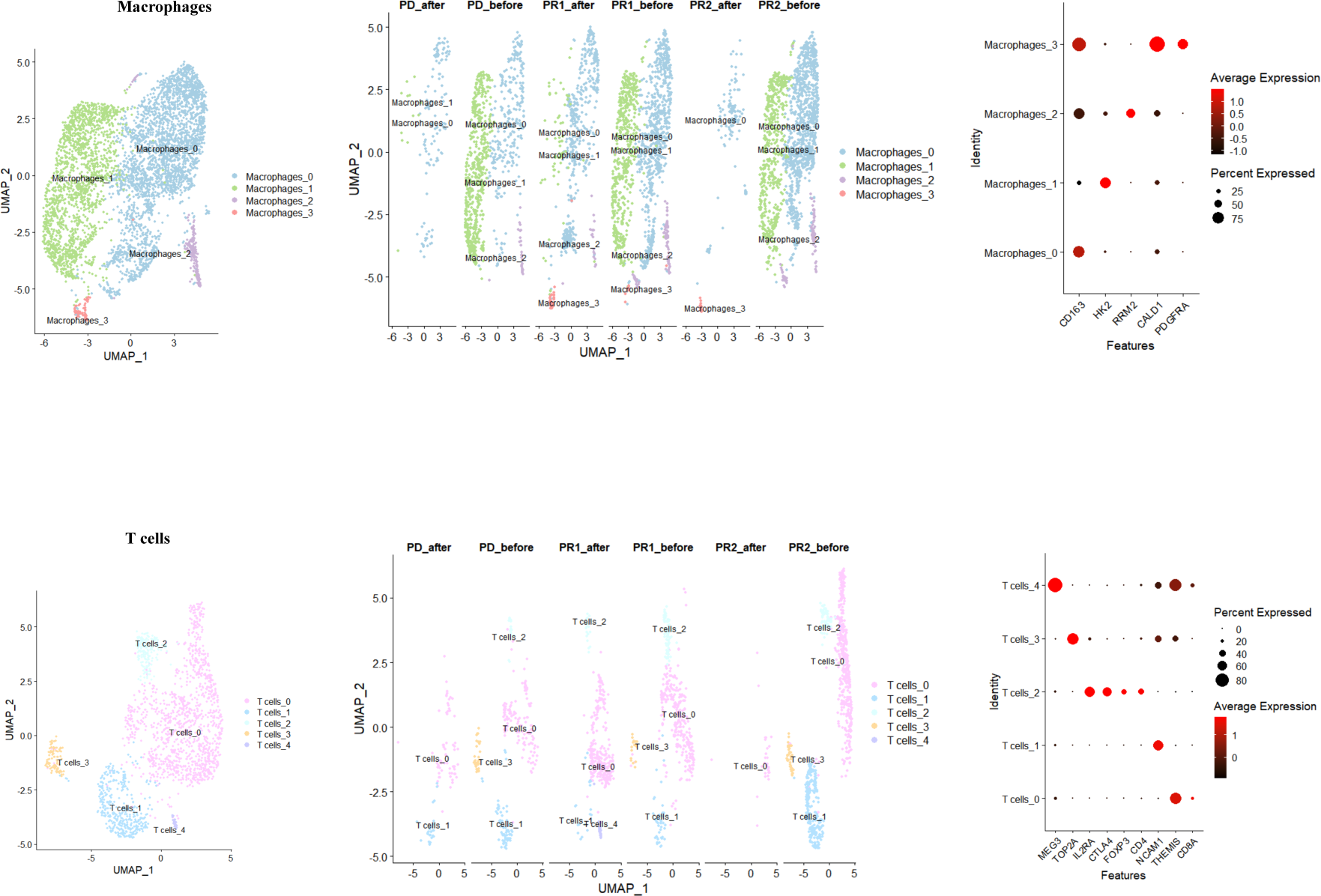
(A) Left: UMAP plot of macrophage subclusters for integrated samples. Middle: UMAP plot of macrophage subclusters for each sample among patients before and after chemotherapy. Right: Dot plot of marker gene expression in each macrophage subcluster. (B) Left: UMAP plot of T cell subclusters for integrated samples. Middle: UMAP plot of T cell subclusters for each sample among patients before and after chemotherapy. Right: Dot plot of marker gene expression in each T cell subcluster. PD_after: Post-chemotherapy sample of the chemoresistant patient. PD_before: Pre-chemotherapy sample of the chemoresistant patient. PR1_after: Post-chemotherapy sample of patient 1 with partial response to chemotherapy. PR1_before: Pre-chemotherapy sample of patient 1 with partial response to chemotherapy. PR2_after: Post-chemotherapy sample of patient 2 with partial response to chemotherapy. PR2_before: Pre-chemotherapy sample of patient 2 with partial response to chemotherapy.

Furthermore, we identified five T cell subclusters (Figure 7), including CD8^+^THEMIS^+^ T cells [21,22] (T cells_0), natural killer (NK) cells (NCAM1^+^CD3^-^) (28) (T cells_1), CD4^+^ regulatory T cells (Tregs) (CD4^+^CTLA4^+^FOXP3^+^IL2RA^+^) (29) (T cells_2), TOP2A^+^ T cells (30) (T cells_3), and MEG3^+^ T cells (31–33) (T cells_4). The CD8^+^ THEMIS^+^ T cells represented a large proportion after treatment. Only 14 genes were found to be overexpressed in the PD_after compared to PR1_after and PR2_after, and no specific pathway enrichment was observed. A small proportion of NKT-like cells remained in the PR1_after and the PD_after, with no DE genes identified between them. These results indicated a generally similar immunosuppressive microenvironment after chemotherapy among patients.

Regarding Lymphatic endothelial cells (Figure 1), only four genes were overexpressed in the PD_after, including RMST, STAT3, NAMPT, and YBX3. For Schwann cells (Figure 1), no DE genes were identified among patients after treatment. Similarly, for Endothelial cells (Figure 1), only 47 DE genes were found to be overexpressed in the PD_after compared to PR1_after and PR2_after, such as NAMPT, SEMA3A, RMST, and ATP1B3, and no significantly enriched pathways were observed. These results indicated that these stromal cells exhibited generally similar characteristics after chemotherapy among patients.

### The NAMPT-INSR was the most prioritized ligand-receptor pair enriched in the chemoresistance patient

Cell-cell interactions play crucial roles in executing cellular functions. Since Epithelia cells_0 cells and Fibroblasts_01_after cells were key cells for chemoresistance, we thus would like to see which cells interacted with them and affected their core genes expression. The genes that were significantly overexpressed in Epithelial cells_0 cells in the PD_after in M3 and M4 modules we identified above were used as the target geneset when Epithelial cells_0 cells were as the receivers. Similarly, the genes that were significantly overexpressed in Fibroblasts_01_after cells in the PD_after in M2 module we identified above were used as the target genesets when Fibroblasts_01_after cells were as the receivers.

Interestingly, results consistently showed that the NAMPT-INSR was the most prioritized ligand-receptor pair for cells interacting with Epithelial cells_0 cells (receiver cells) in the PD_after when compared to PR1_after and PR2_after. For example, INSR was the most potentially interacted receptor for NAMPT for Fibroblasts_01_after cells (sender) interacting with Epithelial cells_0 cells (receiver) in the PD_after compared to PR1_after (Figure 8A left) and PR2_after (Figure 8B left). NAMPT was the only ligand with high ranking for ligand activity, and also with high expression level in Fibroblasts_01_after in the PD_after compared to PR1_after (Figure 8A right) and PR2_after (Figure8B right). The corresponding downstream target genes were IL1R and INSR in the PD_after compared to PR1 (Figure 8A right) and PR2 (Figure 8B right). These results were also applied to other cells interacting with Epithelial cells_0 cells in the PD_after to drive the overexpression of IL1R and INSR as the downstream genes, such as Endothelial cells, Fibroblasts_2, Fibroblasts_3, Lymphatic endothelial cells, and T cells_0, which were given in the Supplementary File 3.

**Figure 8.**
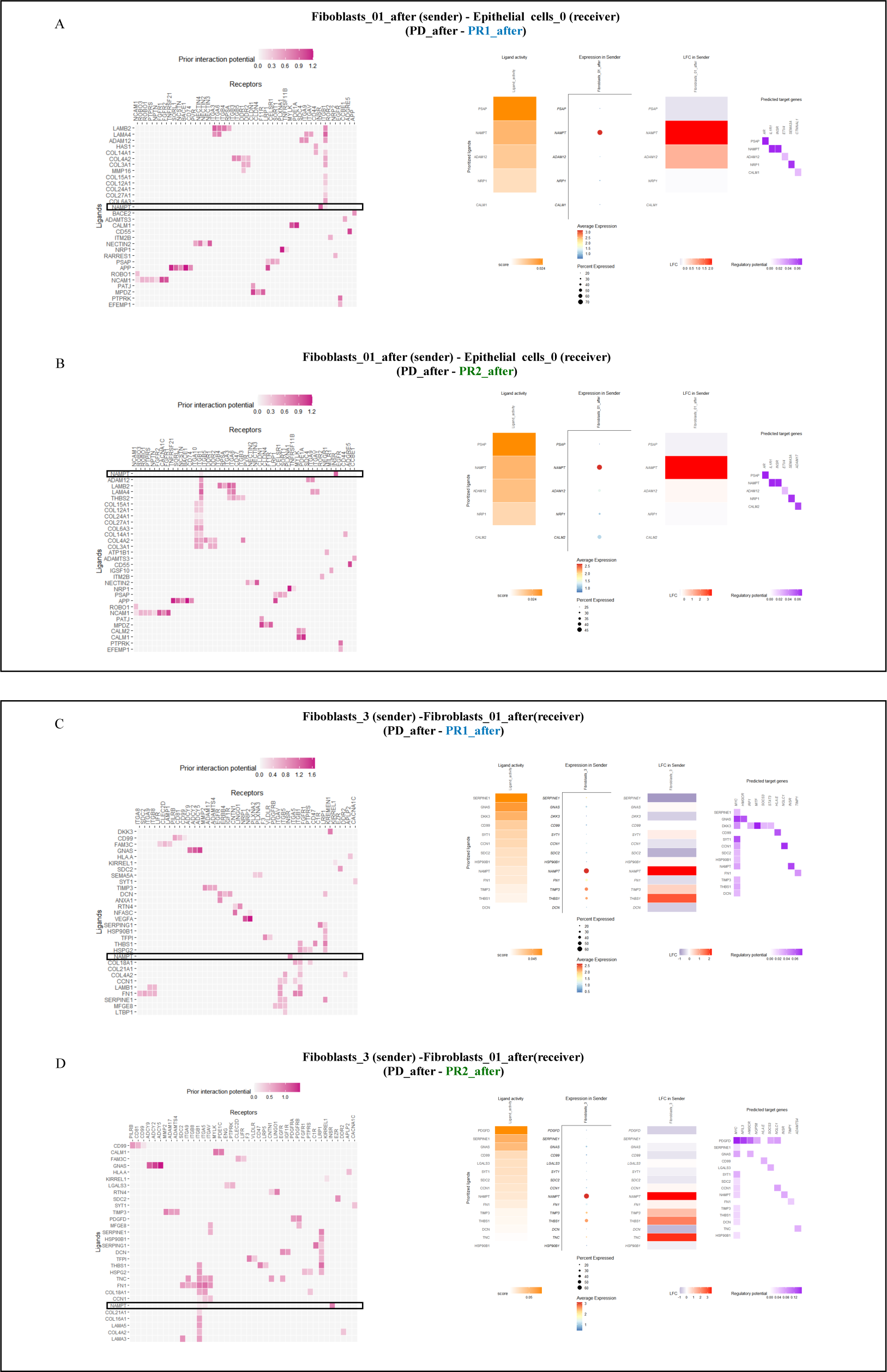
Cell-cell interactions analyses. (A) Ligand-receptor inference (left) and target genes inference (right) when Fibroblasts_01_after cells acted as senders and interacted with Epithelial_cells_0 cells as receivers in the PD_after compared to PR1_after. The NAMPT-INSR was the most prioritized ligand receptor pair, with IL1R and INSR itself as the downstream target genes. (B) Ligand-receptor inference (left) and target genes inference (right) when Fibroblasts_01_after cells acted as senders and interacted with Epithelial_cells_0 cells as receivers in the PD_after compared to PR2_after. The NAMPT-INSR was the most prioritized ligand receptor pair, with IL1R and INSR itself as the downstream target genes. (C) Ligand-receptor inference (left) and target genes inference (right) when Fibroblasts_3 cells acted as senders and interacted with Fibroblasts_01_after cells as receivers in the PD_after compared to PR1_after. The NAMPT-INSR was the most prioritized ligand receptor pair, with MYC and INSR itself as the downstream target genes. (D) Ligand-receptor inference (left) and target genes inference (right) when Fibroblasts_3 cells acted as senders and interacted with Fibroblasts_01_after cells as receivers in the PD_after compared to PR2_after. The NAMPT-INSR was the most prioritized ligand receptor pair, with MYC and INSR itself as the downstream target genes. PD_after: Post-chemotherapy sample of the chemoresistant patient. PR1_after: Post-chemotherapy sample of patient 1 with partial response to chemotherapy. PR2_after: Post-chemotherapy sample of patient 2 with partial response to chemotherapy.

The NAMPT-INSR was also the most prioritized ligand-receptor pair for cells interacting with Fibroblasts_01_after cells (receiver) in the PD_after when compared to PR1_after and PR2_after, with MYC and INSR itself as the downstream target genes, such as Fibroblasts_3 and Lymphatic endothelial cells (Figure 8C and 8D, Supplementary File 3).

## Discussion

In this study, we revealed that chemoresistant cancer cells were pre-existing in individual patients prior to treatment. This observation aligns with existing research that cancer cells are often clustered by patient IDs, highlighting the strong individual differences among cancer cells (34,35), which can contribute to the heterogeneity of tumors in individual patients. These results provide a foundation for developing biomarkers to diagnose chemoresistance before initiating treatment. We identified BMP1 and TPM2 as promising universal candidate biomarkers for identifying chemoresistant patients before treatment when using bulk transcriptome data. Both genes can facilitate chemoresistance (36,37), supporting the rationality of our analyses.

We found two co-expression network modules were signficantly overexpressed in Epithelial cells_0 cells in the chemoresistant sample. One network was formed with hub genes such as ESR1 and CTNNB1. The other module network was formed by tight connections among genes of IGF1R and INSR, AR, NCOR2 and SMAD2. CTNNB1 has been suggested as a potential biomarker for chemoresistance in breast cancer (38), which can be a good candidate therapeutic target for chemoresistance in HGSOC. Notably, two hormone-related genes, ESR1and AR, were involved in these core networks. Hormone imbalance can contribute to the development of ovarian cancer, and both androgen and estrogen have been reported to regulate proliferation and progression in ovarian cancer (39,40). Our findings here thus highlight the critical roles of these hormones in chemoresistance in HGSOC which deserve consideration. Additionally, two hub genes are insulin receptors (IGF1R and INSR). The enrichment of insulin resistance pathway in Epithelial cells_0 cells in the chemoresistant sample further supports the potential link between insulin and early chemoresistance. Insulin is well-known for cellular metabolism and growth (41). It is known that chemotherapy-induced insulin resistance can impair anti-cancer efficacy (42). Elevated circulating insulin due to insulin resistance, coupled with overexpressed insulin receptors in cancer cells can confer a selective advantage to promote proliferation and migration (43,44), and thus contributing to chemoresistance in HGSOC. Our findings thus underscore the importance of insulin-related genes in early chemoresistance in HGSOC.

In addition to cancer cells, other cells in the tumor microenvironment can also play crucial roles in chemotherapy responses. Unlike the pre-existing cancer cells, our findings indicated that the induced fibroblasts (Fibroblasts_01_after) following chemotherapy were key cells associated with chemoresistance. A subnetwork formed by cholesterol biosynthesis-related genes was found in the module that was significantly overexpressed in Fibroblasts_01_after cells in the chemoresistant sample. Cholesterol biosynthesis can support cancer progression and also induce drug resistance (45). For example, gene SREBF2 has been reported to facilitate chemoresistance in ovarian cancer (46). Our results thus implies the importance of cholesterol metabolism in chemoresistance in HGSOC. Another subnetwork was formed by cancer-related genes such as STAT3 and MYC. The overexpression of both genes have well been linked to chemoresistance in ovarian cancer (47,48). Genes within these subnetworks could have critical roles in chemoresistance and are good candidate targets.

Cellular interactions in the tumor microenvironment are vital for chemoresistance. Our analysis revealed that the NAMPT-INSR was the most prioritized ligand-receptor pair enriched in the chemoresistant sample when cells interacting with the two key cell types, i.e. Epithelial cells_0 and Fibroblasts_01_after. NAMPT as a rate-limiting enzyme for NAD synthesis, is required for cellular metabolism and DNA repair, for example, as the substrate of PARPs (a DNA repair enzyme whose inhibitors are widely used in second-line therapy for HGSOC) (49). Recent studies have highlighted the potential function of NAMPT as an alternative ligand for INSR (50), which can enhance insulin resistance (51), which can impair anti-cancer efficacy as we mentioned above. Besides, similar to insulin, NAMPT can stimulate glucose uptake and proliferation through INSR to confer a selective advantage to promote proliferation and migration of cancer cells as we mentioned above. Moreover, NAMPT treatment can induce fibrosis by overproducing profibrotic molecules through the INSR transduction pathway (51). This can contribute to chemoresistance by creating a physical barrier to protect from chemotherapeutic drugs (52). All these findings gave strong supports for the involvement of NAMPT-INSR in chemoresistance in HGSOC patients. We further identified that IL1R1, MYC and INSR were the downstream target genes of NAMPT-INSR. MYC and INSR can induced chemoresistance as we mentioned above. The chemoresistant functions of these target genes gave further support that NAMPT-INSR can have critical roles in chemoresistance in HGSOC patients, although the specific mechnism involved in HGSOC needs further investigation.

Notably, NAMPT was overexpressed in Fibroblasts_01_after cells only after chemotherapy. This aligns with previous findings about the overexpression of NAMPT at the onset of drug resistance (53). NAMPT can be induced by hypoxia via a STAT3-dependent mechanism (54). Additionally, MYC can interact with the NAMPT promoter and stimulate its expression to promote cancer (55,56). The overexpression of both STAT3 and MYC in the PD_after in Fibroblasts_01_after cells in our study can help explain the overexpression of NAMPT in these cells after chemotherapy. Our cellular interactions analyses showed that MYC was also the downstream target gene of NAMPT in Fibroblasts_01_after in the PD_after. Indeed, MYC and NAMPT are involved in a positive feedback loop to drive tumorigenesis (56). The consistent results supported the reliability of our analyses.

The immune microenvironment in patients with varying chemotherapy responses exhibited a general trend of being inhibited and proinflammatory after chemotherapy. The proportions of T cells, both before and after chemotherapy, were observed to be small, which aligns with the characteristic of ovarian cancer being a “cold” tumor (57). Notably, Ciliated-secretory intermediate cells displayed a close transcriptional profile with Epithelial cells_0 cells in the PD_after, consistent with the hypothesis that they are the primary origin of HGSOC, originating from the fallopian tube and fall off to the ovary (24,25). Although other stromal cells showed minimal differences in gene expression among samples after chemotherapy, the majority of them exhibited overexpression of NAMPT in the chemoresistant patient. This is in line with the findings that the NAMPT-INSR pair was highly enriched when these stromal cells interacted with Fibroblasts_01_after cells and Epithelial cells_0 cells in the chemoresistant patient (Supplementary File 3).

## Conclusions

In conclusion, we found that pre-existing cancer cells and induced fibroblasts as key contributors for early chemoresistance in HGSOC patients. Metabolism reprogramming, involving hormone-related genes (ESR1 and AR), insulin-related genes (IGF1R and INSR) and cholesterol biosynthesis-related genes, could play critical roles in early chemoresistant HGSOC. CTNNB1, STAT3 and MYC as core hub genes are also good candidate chemoresistant targets. The NAMPT-INSR ligand-receptor pair enriched for cells interacted with Fibroblasts_01_after cells and Epithelial cells_0 cells, can be important for early chemoresistance in HGSOC.

We thus propose that the combination of chemotherapy with INSR inhibitors and NAMPT inhibitors could represent a promising treatment strategy for early chemoresistant HGSOC patients. The inhibitors of NAMPT and INSR have undergone extensive development and clinical testing (58,59). Previous research has demonstrated that the combination of a NAMPT inhibitor and chemotherapy resulted in a better prognosis than chemotherapy alone in mice with HGSOC (60), holds promise for further improving the therapeutic window. Despite the limitations in sample size, our study has yielded valuable clinic findings, enrich the heterogeneous library of HGSOC and could offer important insights into potential clinical biomarkers and therapeutic targets for HGSOC chemoresistance. The findings from our study may also contribute to advancements in understanding and addressing chemoresistance in various cancer types in a broad sense.

## Data Availability

All data produced in the present study are available upon reasonable request to the authors.

## Acknowledgments

We thank the valuable suggestions from Yunfei Zhao and Prof. Canwei Xia. We also would like to acknowledge the computer support provided by Prof. Yang Liu, Yuqing Han, and Ximin He.

## Authors contributions

Langyu Gu and Guofen Yang proposed the study. Langyu Gu conducted the single nuclei transcriptome data analyses. Shasha He and Yu Zeng conducted chemotherapy responses evaluation. Linxiang Wu proposed the sampling plan and provided the clinical information. Guofen Yang, Hongwei Shen, and Linxiang Wu conducted the sampling. Langyu Gu, Yang Zhang, Chuling Wu, and Huishan Xu conducted survival analyses. Langyu Gu, Chenqing Zheng, and Xiaoyan Zhang conducted pathways enrichment analyses. Guofen Yang, Yufeng Ren, Shasha He, and Shuzhong Yao provided funding supports. Langyu Gu wrote the manuscript, with all authors providing comments and approval.

## Statement of significance

We have identified key cells and core gene networks for early chemoresistance in ovarian cancer.

