## Supplementary File 2 for "Pre-existing cancer cells and induced fibroblasts are key cells for early chemoresistance in ovarian cancer"

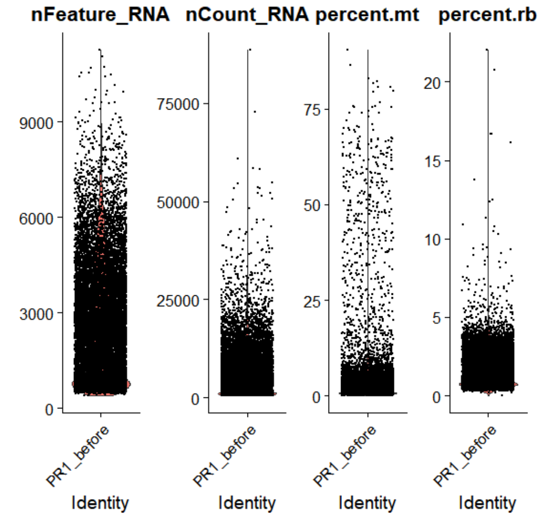


**Figure 1** Violin plots showing QC metrics for PR1_before. nFeature_RNA represents the number of genes detected in each cell, nCount_RNA represents the sum of the expression levels of all genes detected in each cell. percent.mt represents the proportion of the detected mitochondrial genes. percent.rb represents the proportion of the detected ribosomal genes.

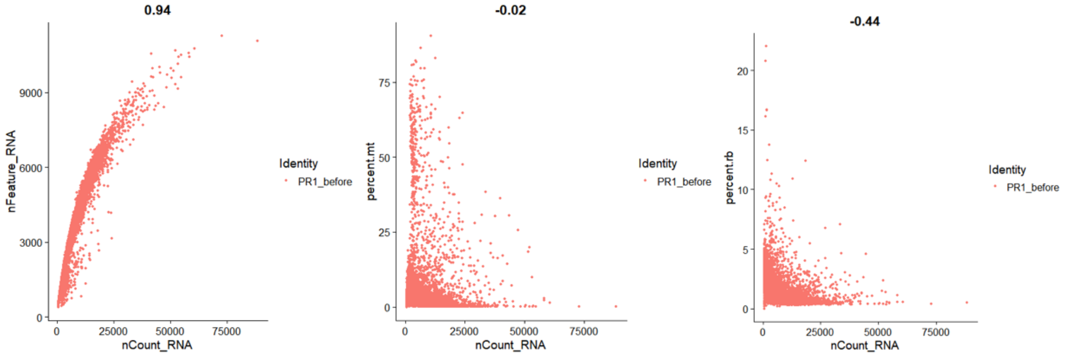


**Figure 2** Scatter plots showing feature-feature relationships in PR1_before. left, scatter plot showing the relationships between nCount_RNA and nFeature_RNA. middle, scatter plot showing the relationships between nCount_RNA and percent.mt. right, scatter plot showing the relationship between nCount_RNA and percent.rb.

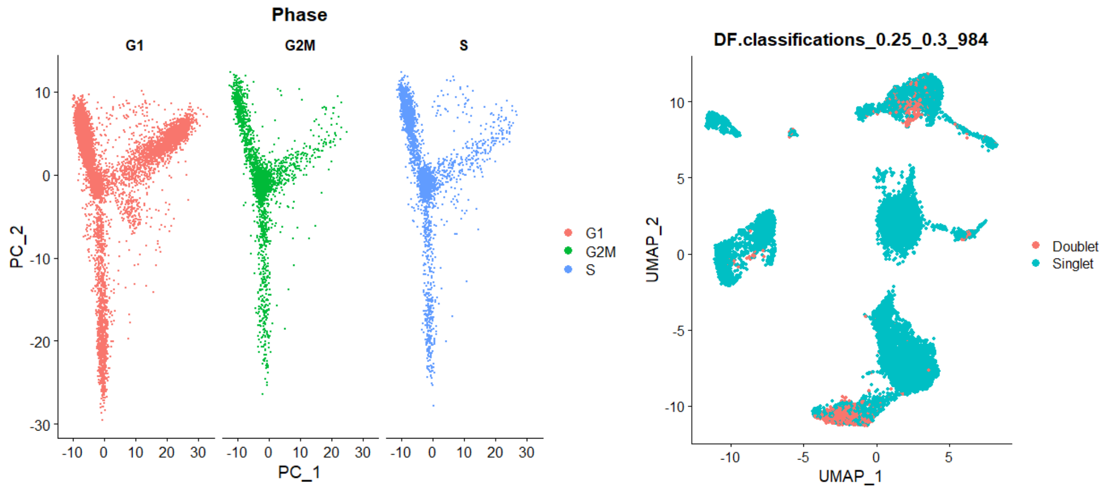


**Figure 3** left, PCA on cell cycle genes revealed that the effects of cell cycle on cells were limited in PR1_before. right, UMAP plot showing the identified doublets in PR1_before.

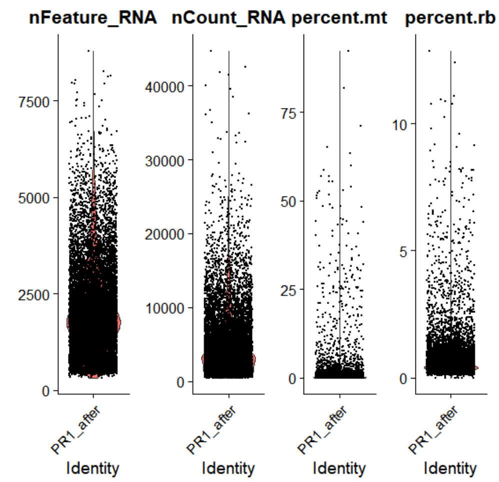


**Figure 4** Violin plots showing QC metrics for PR1_after. nFeature_RNA represents the number of genes detected in each cell, nCount_RNA represents the sum of the expression levels of all genes detected in each cell. percent.mt represents the proportion of the detected mitochondrial genes. percent.rb represents the proportion of the detected ribosomal genes.

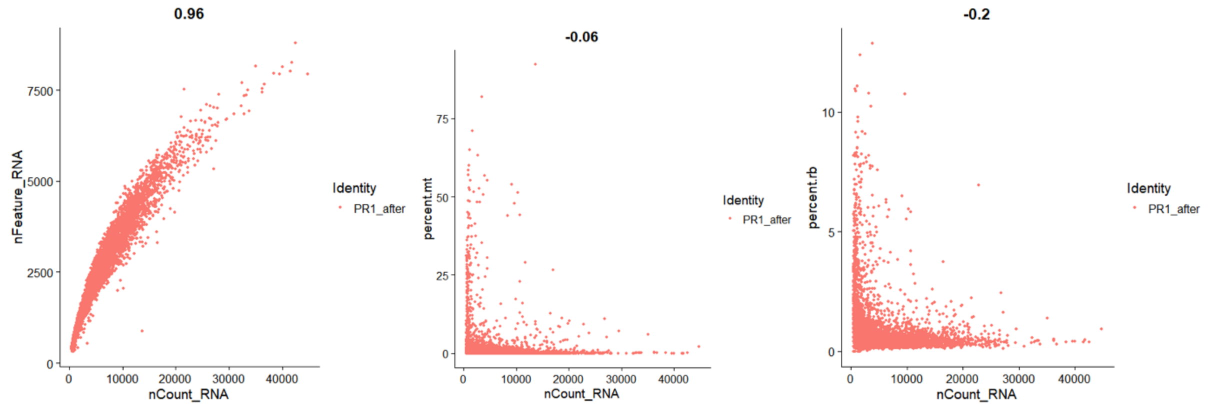


**Figure 5** Scatter plots showing feature-feature relationships in PR1_after. left, scatter plot showing the relationships between nCount_RNA and nFeature_RNA. middle, scatter plot showing the relationships between nCount_RNA and percent.mt. right, scatter plot showing the relationship between nCount_RNA and percent.rb.

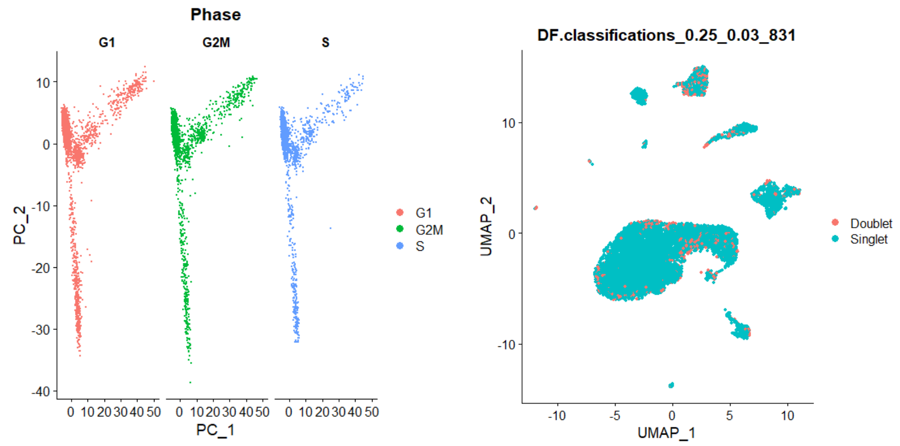


**Figure 6** left, PCA on cell cycle genes revealed that the effects of cell cycle on cells were limited in PR1_after. right, UMAP plot showing the identified doublets in PR1_after.

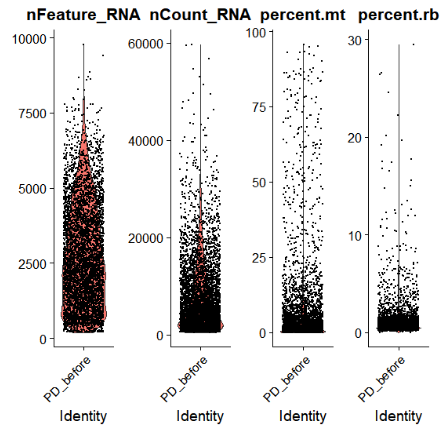


**Figure 7** Violin plots showing QC metrics for PD_before. nFeature_RNA represents the number of genes detected in each cell, nCount_RNA represents the sum of the expression levels of all genes detected in each cell. percent.mt represents the proportion of the detected mitochondrial genes. percent.rb represents the proportion of the detected ribosomal genes.

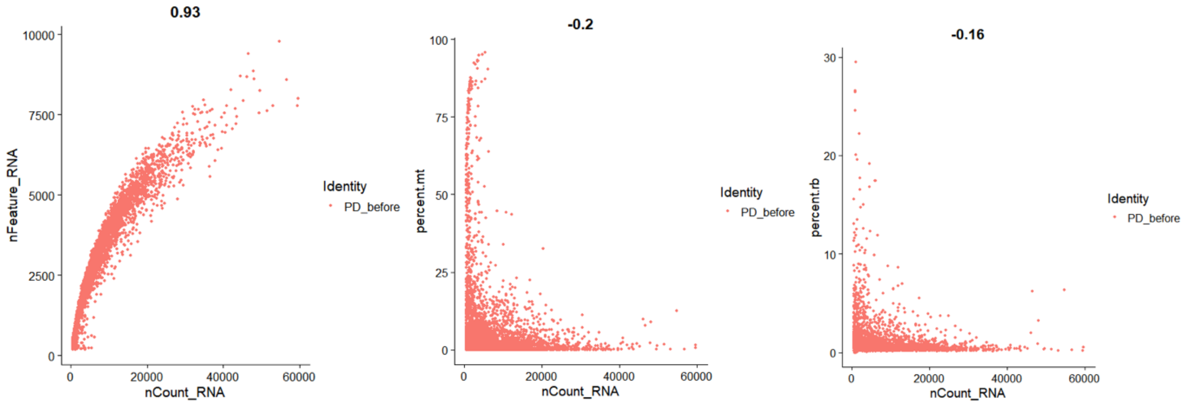


**Figure 8** Scatter plots showing feature-feature relationships in PD_before. left, scatter plot showing the relationships between nCount_RNA and nFeature_RNA. middle, scatter plot showing the relationships between nCount_RNA and percent.mt. right, scatter plot showing the relationship between nCount_RNA and percent.rb.

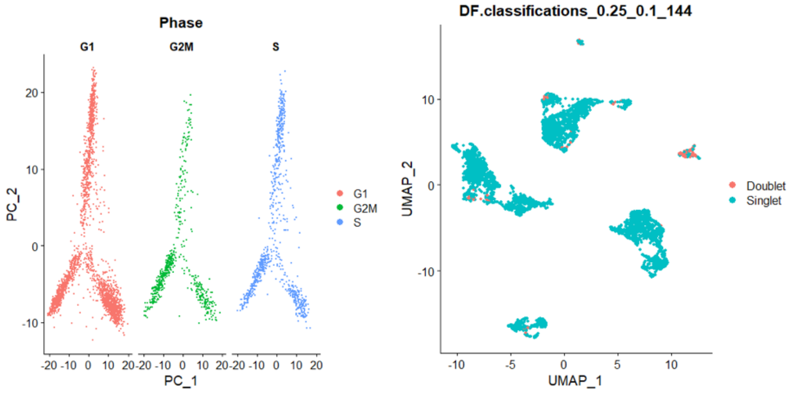


**Figure 9** left, PCA on cell cycle genes revealed that the effects of cell cycle on cells were limited in PD_before. right, UMAP plot showing the identified doublets in PD_before.

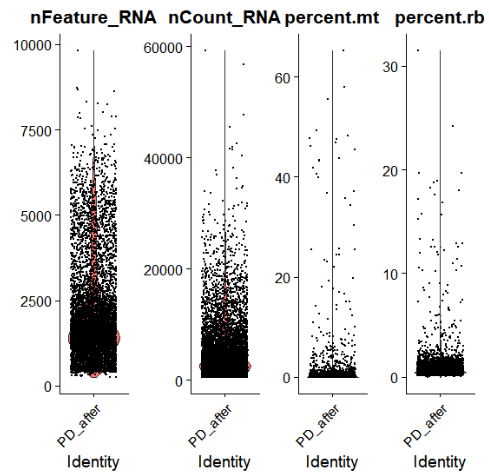

**Figure 10** Violin plots showing QC metrics for PD_after. nFeature_RNA represents the number of genes detected in each cell, nCount_RNA represents the sum of the expression levels of all genes detected in each cell. percent.mt represents the proportion of the detected mitochondrial genes. percent.rb represents the proportion of the detected ribosomal genes.

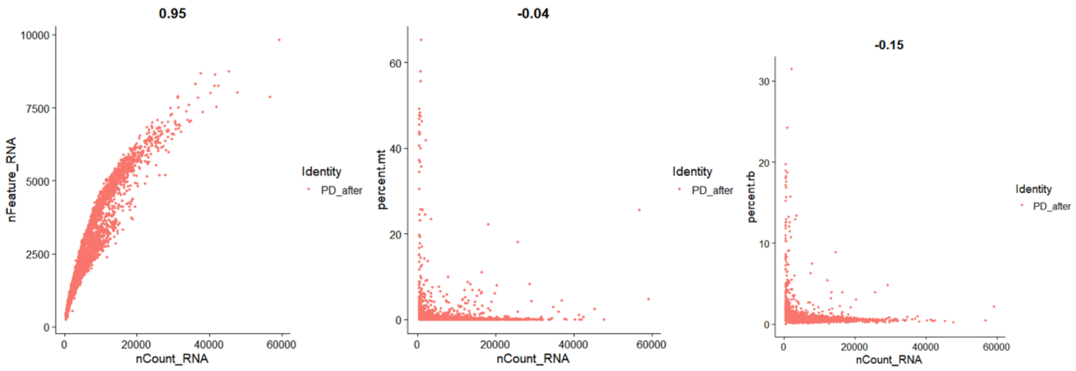

**Figure 11** Scatter plots showing feature-feature relationships in PD_after. left, scatter plot showing the relationships between nCount_RNA and nFeature_RNA. middle, scatter plot showing the relationships between nCount_RNA and percent.mt. right, scatter plot showing the relationship between nCount_RNA and percent.rb.

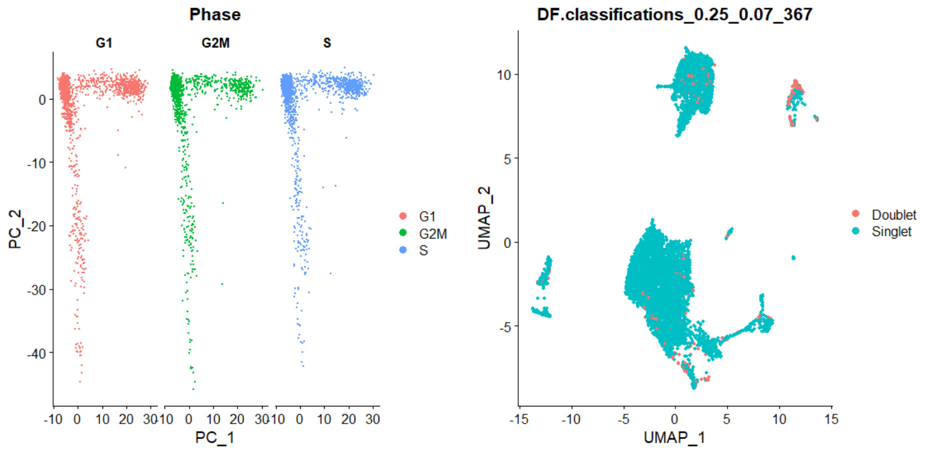

**Figure 12** left, PCA on cell cycle genes revealed that the effects of cell cycle on cells were limited in PD_after. right, UMAP plot showing the identified doublets in PD_after.

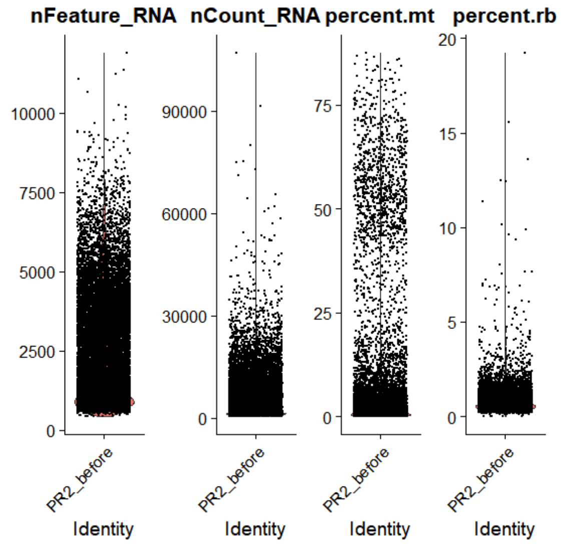


**Figure 13** Violin plots showing QC metrics for PR2_before. nFeature_RNA represents the number of genes detected in each cell, nCount_RNA represents the sum of the expression levels of all genes detected in each cell. percent.mt represents the proportion of the detected mitochondrial genes. percent.rb represents the proportion of the detected ribosomal genes.

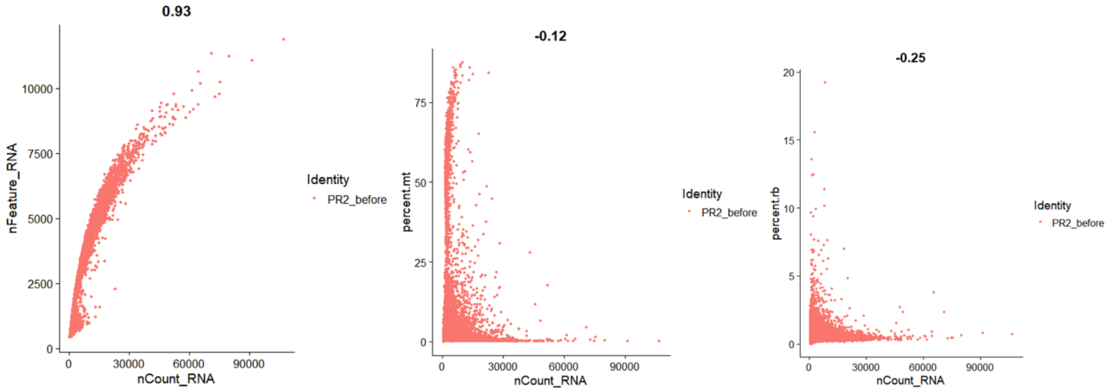


**Figure 14** Scatter plots showing feature-feature relationships in PR2_before. left, scatter plot showing the relationships between nCount_RNA and nFeature_RNA. middle, scatter plot showing the relationships between nCount_RNA and percent.mt. right, scatter plot showing the relationship between nCount_RNA and percent.rb.

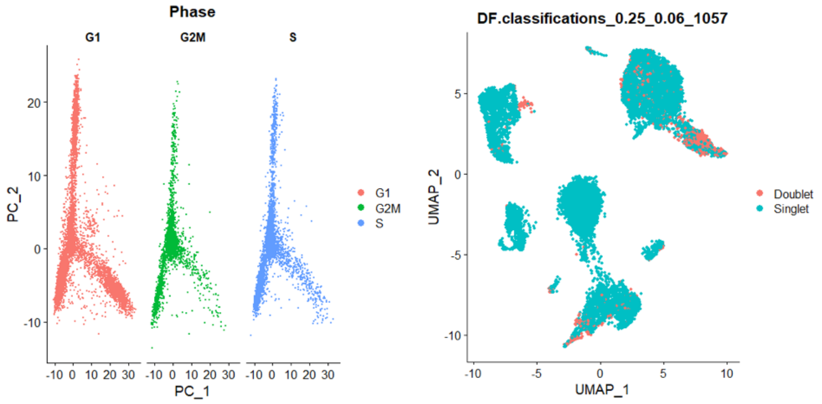


**Figure 15** left, PCA on cell cycle genes revealed that the effects of cell cycle on cells were limited in PR2_before. right, UMAP plot showing the identified doublets in PR2_before.

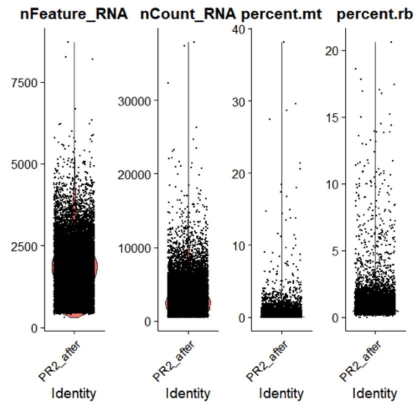


**Figure 16** Violin plots showing QC metrics for PR2_after. nFeature_RNA represents the number of genes detected in each cell, nCount_RNA represents the sum of the expression levels of all genes detected in each cell. percent.mt represents the proportion of the detected mitochondrial genes. percent.rb represents the proportion of the detected ribosomal genes.

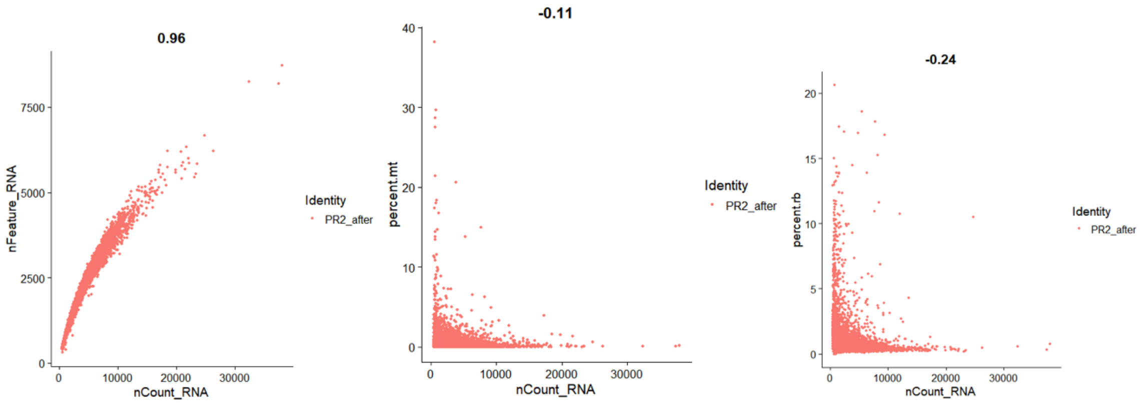


**Figure 17** Scatter plots showing feature-feature relationships in PR2_after. left, scatter plot showing the relationships between nCount_RNA and nFeature_RNA. middle, scatter plot showing the relationships between nCount_RNA and percent.mt. right, scatter plot showing the relationship between nCount_RNA and percent.rb.

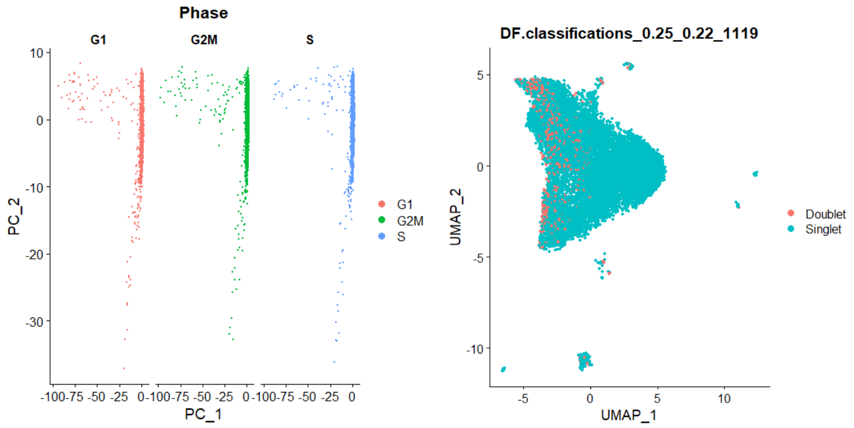


**Figure 18** left, PCA on cell cycle genes revealed that the effects of cell cycle on cells were limited in PR2_after. right, UMAP plot showing the identified doublets in PR2_after.
