## Supplementary File 3 for "Pre-existing cancer cells and induced fibroblasts are key cells for early chemoresistance in ovarian cancer"

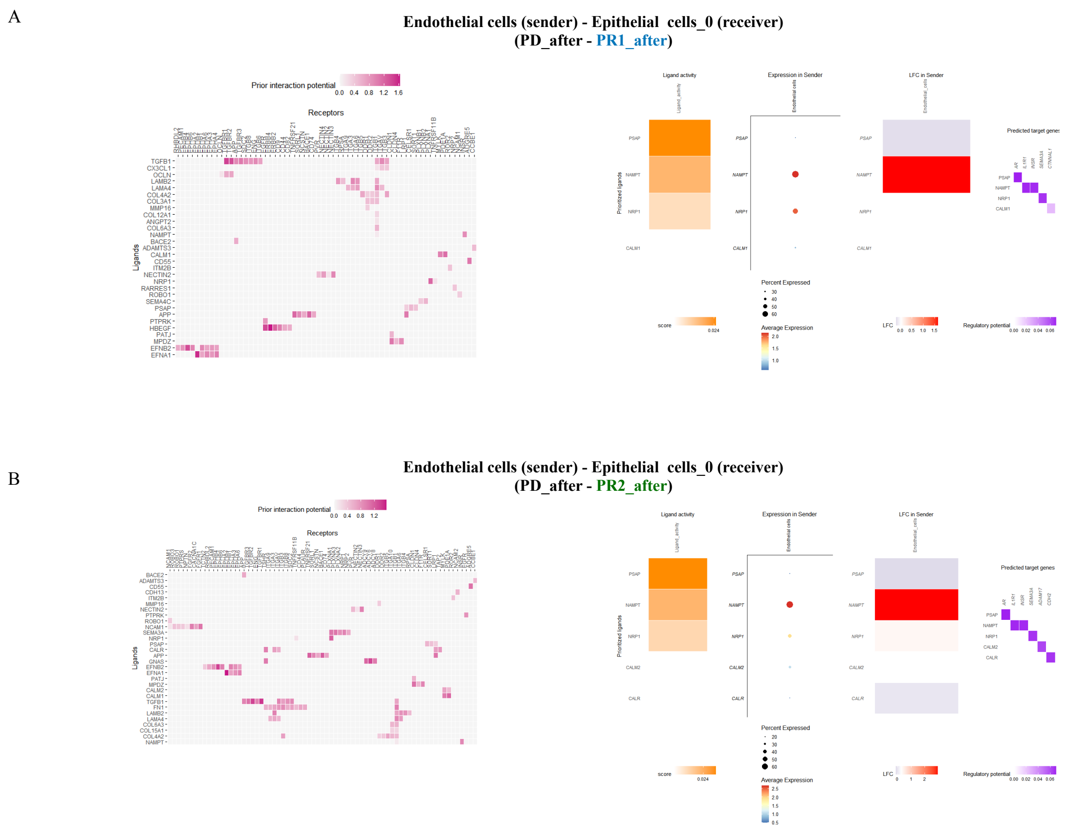


**Figure 1** Cell-cell interactions analyses. (A) Ligand-receptor inference (left) and target genes inference (right) when Endothelial cells were as senders interacted with Epithelial_cells_0 cells (receivers) in the PD_after when compared to PR1_after. (B) Ligand-receptor inference (left) and target genes inference (right) when Endothelial cells were as senders interacted with Epithelial_cells_0 cells (receivers) in the PD_after when compared to PR2_after. PD_after: Post-chemotherapy sample of the chemoresistant patient. PR1_after: Post-chemotherapy sample of patient 1 with partial response to chemotherapy. PR2_after: Post-chemotherapy sample of patient 2 with partial response to chemotherapy.

A

**Fibroblasts_2 (sender) - Epithelial cells_0 (receiver)**
**(PD_after - PR1_after)**

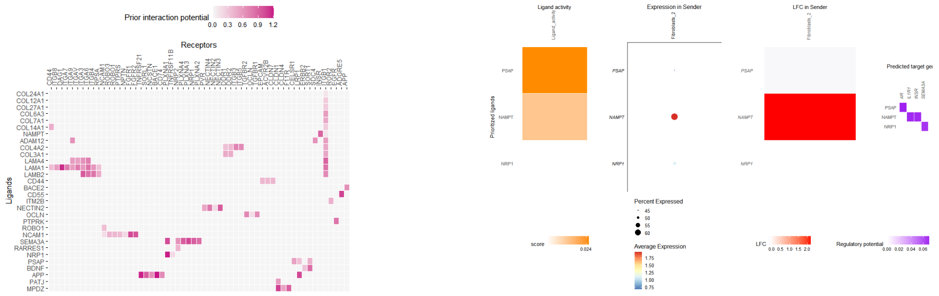


B

**Fibroblasts_2 (sender) - Epithelial cells_0 (receiver)**
**(PD_after - PR2_after)**

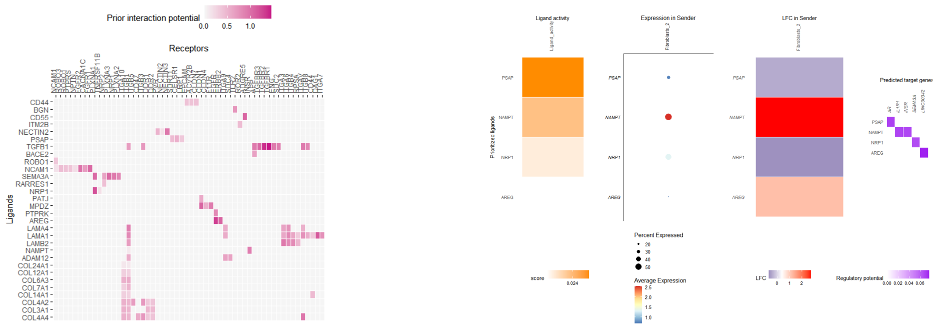


**Figure 2** Cell-cell interactions analyses. (A) Ligand-receptor inference (left) and target genes inference (right) when Fibroblastss_2 cells were as senders interacted with Epithelial_cells_0 cells (receivers) in the PD_after when compared to PR1_after. (B) Ligand-receptor inference (left) and target genes inference (right) when Fibroblasts_2 cells were as senders interacted with Epithelial_cells_0 cells (receivers) in PD_after when compared to PR2_after. PD_after: Post-chemotherapy sample of the chemoresistant patient. PR1_after: Post-chemotherapy sample of patient 1 with partial response to chemotherapy. PR2_after: Post-chemotherapy sample of patient 2 with partial response to chemotherapy.

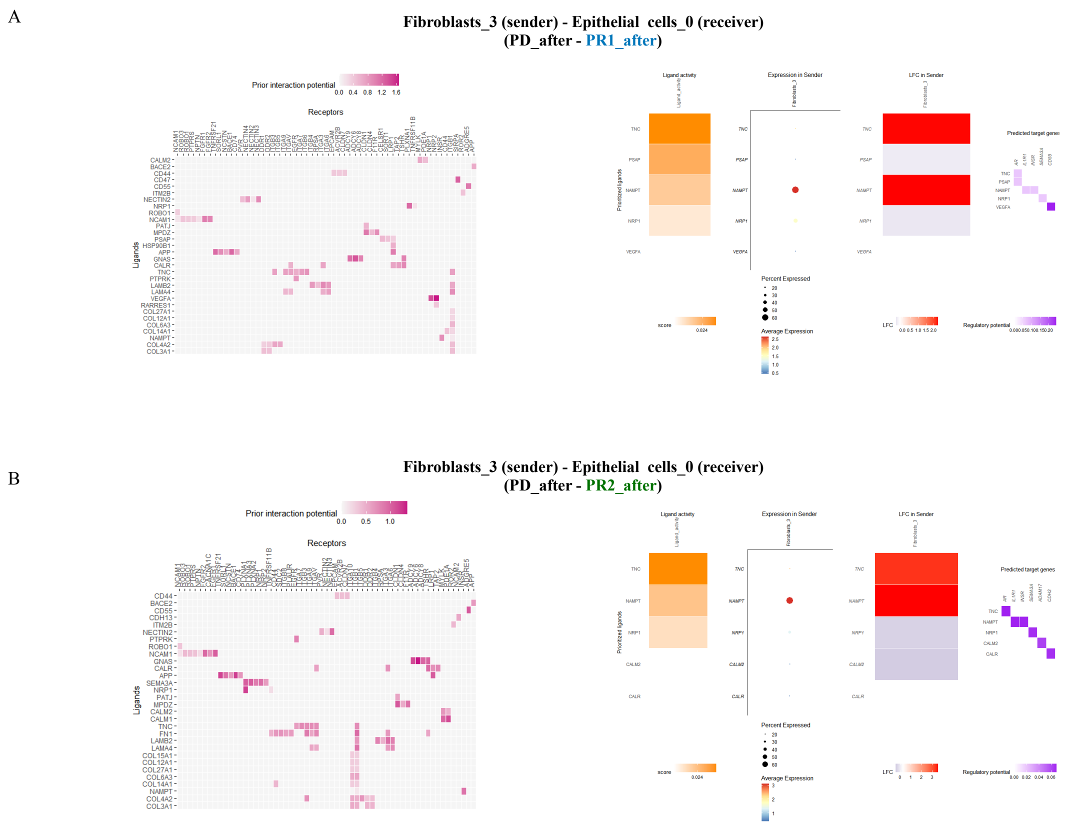


**Figure 3** Cell-cell interactions analyses. (A) Ligand-receptor inference (left) and target genes inference (right) when Fibroblastss_3 cells were as senders interacted with Epithelial_cells_0 cells (receivers) in the PD_after when compared to PR1_after. (B) Ligand-receptor inference (left) and target genes inference (right) when Fibroblasts_3 cells were as senders interacted with Epithelial_cells_0 cells (receivers) in the PD_after when compared to PR2_after. PD_after: Post-chemotherapy sample of the chemoresistant patient. PR1_after: Post-chemotherapy sample of patient 1 with partial response to chemotherapy. PR2_after: Post-chemotherapy sample of patient 2 with partial response to chemotherapy.

A

**Lymphatic endothelial cells (sender) - Epithelial cells_0 (receiver)**
**(PD_after - PR1_after)**

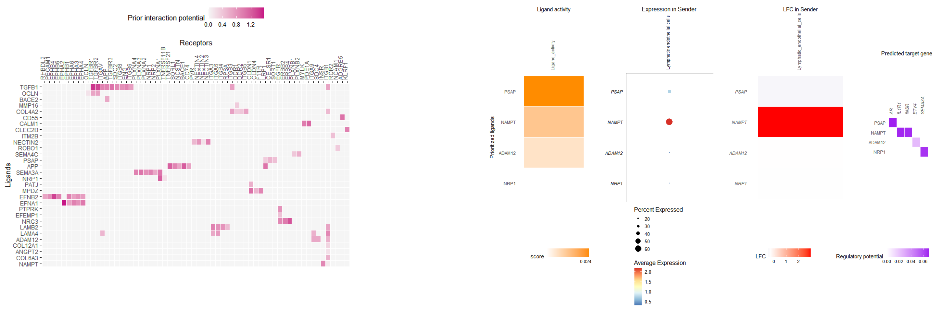


B

**Lymphatic endothelial cells (sender) - Epithelial cells_0 (receiver)**
**(PD_after - PR2_after)**

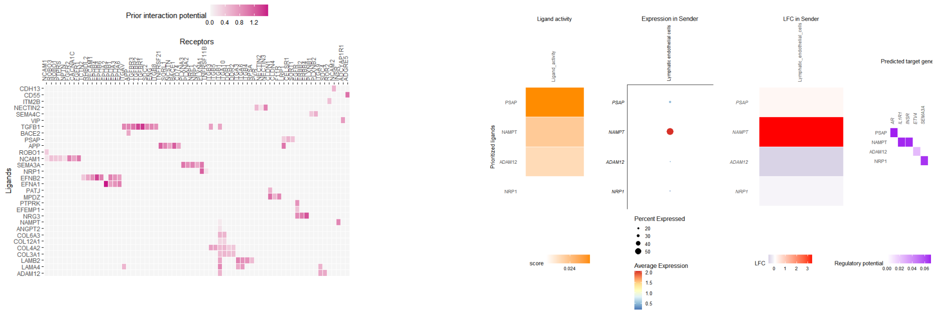


**Figure 4** Cell-cell interactions analyses. (A) Ligand-receptor inference (left) and target genes inference (right) when Lymphatic endothelial cells were as senders interacted with Epithelial_cells_0 cells (receivers) in the PD_after when compared to PR1_after. (B) Ligand-receptor inference (left) and target genes inference (right) when Lymphatic endothelial cells were as senders interacted with Epithelial_cells_0 cells (receivers) in the PD_after when compared to PR2_after. PD_after: Post-chemotherapy sample of the chemoresistant patient. PR1_after: Post-chemotherapy sample of patient 1 with partial response to chemotherapy. PR2_after: Post-chemotherapy sample of patient 2 with partial response to chemotherapy.

A

T cells_0 (sender) - Epithelial cells_0 (receiver)
(PD_after - PR1_after)

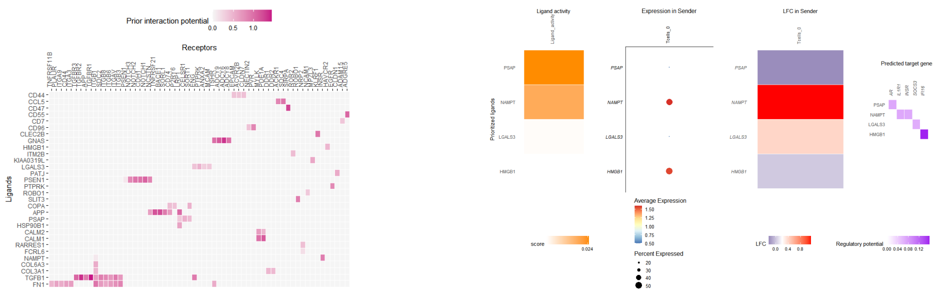


B

T cells_0 (sender) - Epithelial cells_0 (receiver)
(PD_after - PR2_after)

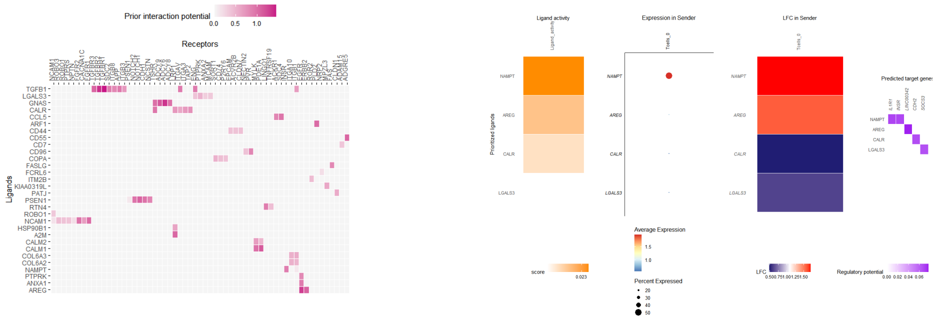


**Figure 5** Cell-cell interactions analyses. (A) Ligand-receptor inference (left) and target genes inference (right) when T cells_0 cells were as senders interacted with Epithelial_cells_0 cells (receivers) in the PD_after when compared to PR1_after. (B) Ligand-receptor inference (left) and target genes inference (right) when T cells_0 cells were as senders interacted with Epithelial_cells_0 cells (receivers) in the PD_after when compared to PR2_after. PD_after: Post-chemotherapy sample of the chemoresistant patient. PR1_after: Post-chemotherapy sample of patient 1 with partial response to chemotherapy. PR2_after: Post-chemotherapy sample of patient 2 with partial response to chemotherapy.

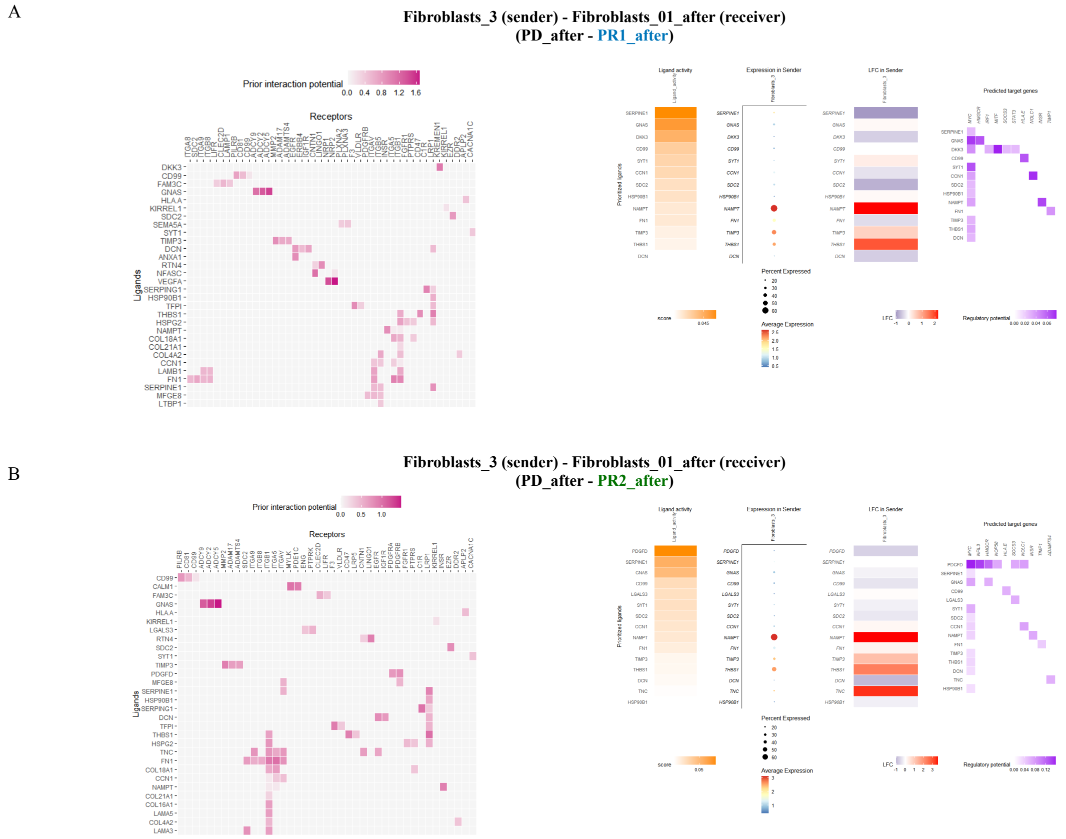


**Figure 6** Cell-cell interactions analyses. (A) Ligand-receptor inference (left) and target genes inference (right) when Fibroblasts_3 cells were as senders interacted with Fibroblasts_01_after cells (receivers) in the PD_after when compared to PR1_after. (B) Ligand-receptor inference (left) and target genes inference (right) when Fibroblasts_3 cells were as senders interacted with Fibroblasts_01_after cells (receivers) in the PD_after when compared to PR2_after. PD_after: Post-chemotherapy sample of the chemoresistant patient. PR1_after: Post-chemotherapy sample of patient 1 with partial response to chemotherapy. PR2_after: Post-chemotherapy sample of patient 2 with partial response to chemotherapy.

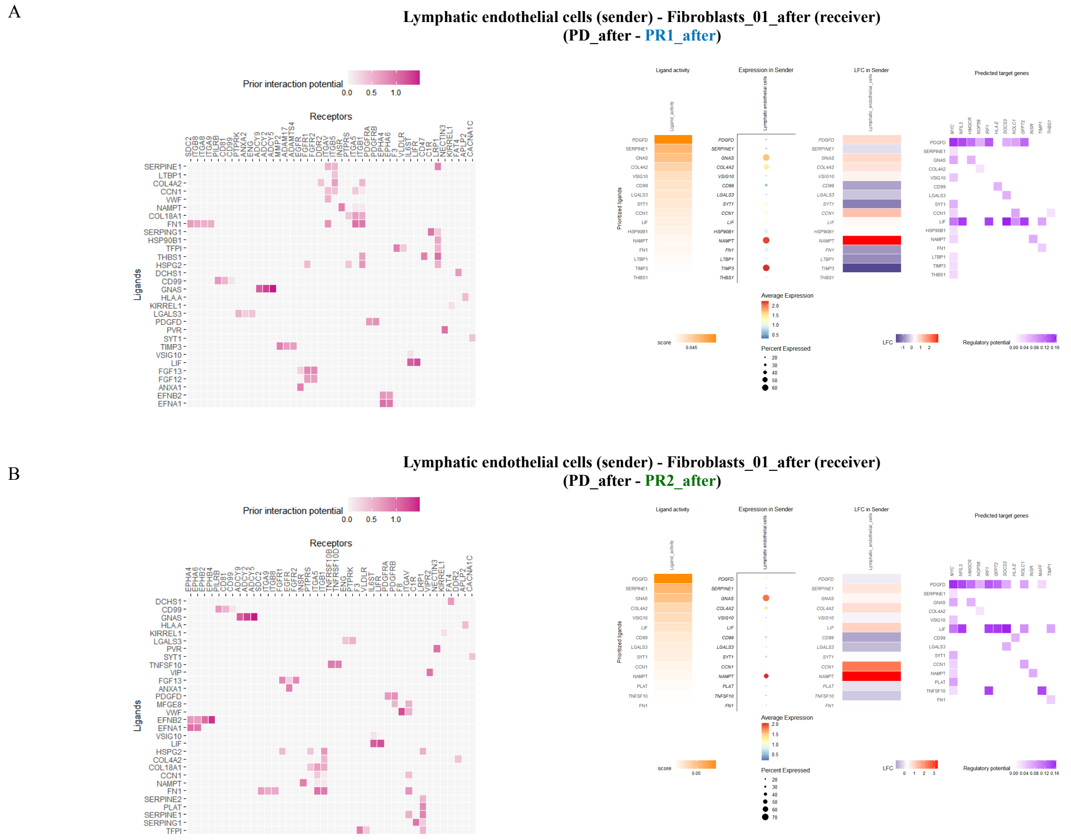


**Figure 7** Cell-cell interactions analyses. (A) Ligand-receptor inference (left) and target genes inference (right) when Lymphatic endothelial cells were as senders interacted with Fibroblasts_01_after cells (receivers) in the chemoresistant sample (PD_after) when compared to PR1_after. (B) Ligand-receptor inference (left) and target genes inference (right) when Lymphatic endothelial cells were as senders interacted with Fibroblasts_01_after cells (receivers) in the chemoresistant sample (PD_after) when compared to PR2_after. PD_after: Post-chemotherapy sample of the chemoresistant patient. PR1_after: Post-chemotherapy sample of patient 1 with partial response to chemotherapy. PR2_after: Post-chemotherapy sample of patient 2 with partial response to chemotherapy.
